# Multiplexed detection, partitioning, and persistence of wild type and vaccine strains of measles, mumps, and rubella viruses in wastewater

**DOI:** 10.1101/2024.05.23.24307763

**Authors:** Jingjing Wu, Michael X. Wang, Prashant Kalvapalle, Michael Nute, Todd J. Treangen, Katherine Ensor, Loren Hopkins, Rachel Poretsky, Lauren B. Stadler

## Abstract

Wastewater surveillance of vaccine-preventable diseases may provide early warning of outbreaks and identify areas to target for immunization. To advance wastewater monitoring of measles, mumps, and rubella viruses, we developed and validated a multiplexed RT-ddPCR assay for the detection of their RNA. Because the measles-mumps-rubella (MMR) vaccine is an attenuated live virus vaccine, we also developed an assay that distinguishes between wild-type and vaccine strains of measles in wastewater and validated it using a wastewater sample collected from a facility with an active measles outbreak. We also evaluated the partitioning behavior the viruses in between the liquid and solid fractions of influent wastewater. We found that assaying the liquid fraction of the wastewater resulted in more sensitive detection of the viruses despite the fact that the viral RNA was enriched in the solid fraction due to the low solids content of the influent wastewater. Finally, we investigated the stability of measles, mumps, and rubella RNA in wastewater samples spiked with viruses over 28 days at two different concentrations and two temperatures (4°C and room temperature) and observed limited viral decay. Our study supports the feasibility of wastewater monitoring for measles, mumps, and rubella viruses for population-level surveillance.

## 1. Introduction

Wastewater monitoring is a powerful tool for infectious disease surveillance that has been used to inform targeted public health interventions. Numerous use cases for how wastewater monitoring has been used by public health officials for decision-making have been demonstrated, including as an early warning system that can indicate disease trends and hospitalizations before other surveillance data (e.g. clinical testing) ^1–7^, as a source of information on disease trends where there is limited case data ^6,8–11^, and as a tool for detecting localized outbreaks within communities ^12–21^ and facilities ^22–31^. Another important use case for wastewater monitoring is to detect vaccine-preventable diseases and direct immunization resources. For example, SARS-CoV-2 and influenza wastewater monitoring at preK-12 schools has been used to decide where and when to provide community immunization events ^31^. Another example is the use of wastewater monitoring for poliovirus as part of the Global Polio Eradication Initiative to inform polio eradication efforts.

Measles is an extremely contagious virus that is vaccine-preventable. It often presents as a rash and can result in complications in unvaccinated individuals including encephalitis and death. In 2022, measles infections caused an estimated 136,000 deaths worldwide, primarily unvaccinated and under vaccinated children ^32^. In the U.S., measles was declared eliminated in 2000, thanks to a highly effective vaccination program. However, measles cases in the U.S. are on the rise, and in 2024 alone, as of April, 2024 a total of 125 cases have been reported in 18 states. Vaccination rates for measles, administered as the measles-mumps-rubella (MMR) vaccine, have decreased among school-aged children below the 95% coverage rate needed for herd immunity. The Centers for Disease Control and Prevention (CDC) estimates that over 61 million MMR vaccine doses were postponed or missed between 2020 and 2022 due to COVID-19 delays. With measles activity on the rise globally, cases arising from contact with an unvaccinated person abroad or visitors can result in transmission in under-vaccinated populations in the US.

Wastewater monitoring has been proposed as a promising tool for population-level monitoring of measles, mumps, and rubella, but methods for their detection and knowledge of their stability in wastewater are limited. Previous studies have shown that measles, mumps, and rubella were detected in urine of infected patients ^33–36^, and measles and rubella can be detected in wastewater ^37–40^. One challenge in measles, mumps, and rubella wastewater detection is that the MMR vaccine is an attenuated (weakened) live virus vaccine. Vaccinated individuals also shed vaccine virus in their urine ^41,42^, and the measles virus has been detected in urine between 1 and 14 days after vaccination in healthy children ^43^. Thus, methods are needed that can distinguish between wild type and vaccine strains in order to differentiate between an outbreak and vaccination event. Further, research is needed to understand the fate of measles, mumps, and rubella viruses and their genetic markers in wastewater, including understanding their partitioning behavior (solid/liquid distribution) and decay in wastewater matrices. This information is critical for optimizing wastewater sampling and storage, as well as laboratory detection methods.

In this study, we developed and validated a multiplexed reverse transcription-droplet digital PCR (RT-ddPCR) assay for the detection of measles, mumps, and rubella viral RNA in wastewater. For measles, we also developed and validated assays that distinguish between wild-type strains (genotypes D8 and B3, which are the current dominant circulating lineages) and vaccine strains. We performed experiments to evaluate the partitioning and concentration of the viruses in the liquid versus solid fraction of influent wastewater samples. Finally, we characterized the persistence of measles, mumps, and rubella RNA in wastewater samples spiked with viruses over 28 days at two different concentrations and two temperatures (4°C and room temperature).

## 2. Materials and Methods

### 2.1. Assay development for measles, mumps and rubella viruses

We designed two different assays for wild type measles (WT1 and WT2), an assay for the measles vaccine strain (VA), an assay for mumps virus, and an assay for rubella virus. Assay details for measles, mumps, and rubella are provided in SI 1.1. All the measles assays use the same forward and reverse primers, but different probe sequences. The mumps and rubella virus assays do not distinguish between wild type and vaccine strains. For measles virus, the assay design focused on the detection of the dominant measles genotypes (D8 and B3) of the current outbreak. We multiplexed the WT1, mumps, and rubella assays and validated them (section 2.3). We also compared WT1, WT2, and VA assays in their ability to distinguish between wild type and vaccine strains of measles. Compared to the WT1 probe, which has only one mismatch to the Edmonston vaccine strains, the WT2 probe sequence has 2 to 3 mismatches to vaccine strains. The WT2 probe also perfectly matches the D8 strain and has only 1 mismatch to the B3 strain (Table SI.8). We duplexed the measles WT2 and VA assays and validated them as described in section 2.3.

For measles, we also designed primers for Sanger sequencing based on the same multiple sequence alignment (MSA) described in SI 1.1. A consensus sequence was generated within a conserved region at the head of the L gene and the primers were designed with Primer3Plus (settings in SI 1.1). To analyze measles Sanger sequencing results, a consensus sequence was generated based on the output sequences of 6 Sanger sequencing replicates (3 forward and 3 reverse). The consensus was then input to NCBI BLAST+ ^44^ (version 2.15.0, blastn was selected for search algorithm, other parameters were kept as default) against all measles sequences (NCBI taxonomy ID 11234) in the nucleotide collection (nt/nr) database (version Apr. 25, 2024), and 822 hits were found. An MSA was generated with MAFFT ^45^ (version 7, default parameters), based on selected BLAST hits to represent each measles genotype, as well as the Sanger consensus. A phylogenetic tree was generated and plotted with iTOL ^46^.

### 2.2. Wastewater sample collection and target viral RNA quantification

Composite influent wastewater samples were collected using a refrigerated time-weighted autosampler from the influent channel of wastewater treatment plants (WWTPs) in Houston between March 4 and April 15, 2024 as described previously ^47^. Influent samples from the top 10 largest WWTPs (by population) were mixed together and used for assay validation, evaluation of the recovery rates of measles, mumps, and rubella viral RNA in wastewater liquid and solid fractions, and viral RNA persistence experiments.

Grab samples collected from a manhole located outside of a facility with an ongoing outbreak were used for measles assay validation. The first confirmed case in the facility was identified on March 7, 2024 and the wastewater samples were collected on March 22 and March 30, 2024. The wastewater samples were frozen at −20°C before being shipped overnight to Rice University on ice. Samples were stored at 4°C upon receipt and processed within 24 hours.

Wastewater liquid and solid fractions were separated and concentrated as described in SI 1.2. The nucleic acid extraction was performed using Chemagic^TM^ Prime Viral DNA/RNA 300 Kit H96 (Chemagic, CMG-1433, PerkinElmer) and the concentrations of target viral RNA were quantified using one-step RT-ddPCR Advanced Kit for Probes (1864021, Bio-Rad) (detailed in SI 1.2). Primer and probe sequences for each assay used in this study, RT-ddPCR reaction assay composition, and thermal cycler conditions are provided in Tables SI.1-4. We followed the EMMI Guidelines ^48^ for quality control and additional information is provided in SI 1.8.

### 2.3. Measles, mumps, and rubella RT-ddPCR assay validation

We validated the measles, mumps, and rubella RT-ddPCR assays using synthetic gblock DNA gene fragments (Integrated DNA Technologies, Coralville, LA, USA), and then using ATCC viral standards, including the Edmonston strain measles virus (VR-24, ATCC), B3 strain measles virus (VR-1981, ATCC), mumps virus (VR-106, ATCC), and rubella virus (VR-1359, ATCC) (Figure SI.1). The ATCC viral standards were heat-lysed at 95°C for 10 minutes using C1000 Touch™ Thermal Cycler (1851197, Bio-Rad) before quantification via RT-ddPCR. Assays were performed in singleplex and multiplex to ensure there were no interactions between primers and probes that would impact quantification results (Figure SI.2 and Table SI.9). A t-test was used to compare the viral RNA concentrations determined using the singleplex and multiplex assays.

To further validate the multiplexed measles (WT1), mumps, and rubella assay, a mixture of the Edmonston strain measles virus, mumps virus, and rubella virus standards were serially diluted and spiked into 50 mL of influent wastewater. In addition, we also verified that the measles WT1, WT2, and VA assays could distinguish between wild-type and vaccine strains in wastewater using wastewater samples spiked with the B3 and Edmonston measles strains. For this experiment, we compared (1) unspiked wastewater samples, (2) wastewater spiked with B3 strain, (3) wastewater spiked with Edmonston strain, and (4) wastewater spiked with both B3 and Edmonston strains. Each condition was performed in triplicate. The liquid fraction of the spiked wastewater samples was concentrated using HA filtration, followed by RNA extraction, and target RNA quantification as described in Section 2.2. We also validated the measles WT2 and VA assays using wastewater samples collected from a manhole outside of a facility with a confirmed outbreak of measles. These samples were shipped from Chicago, IL to Rice University and processed as described above.

Amplification of a 437 bp region of the measles genome was performed and sequenced for confirmation and genotyping. Briefly, the RNA extracts from select measles-positive facility wastewater samples were used as template in RT-PCR using the SuperScript™ IV One-Step RT-PCR System (12594025, Invitrogen) with primers for Sanger sequencing (Table SI.5) and following the manufacturer’s protocol. RT-PCR reaction assay composition and thermal cycler conditions are provided in Tables SI.6-7. The RT-PCR products were visualized with gel electrophoresis and purified using the Monarch DNA gel extraction kit (T1020S, NEB) following the manufacturer’s instructions. The purified cDNA was diluted to 1 ng/µL and submitted for Sanger sequencing (GeneWiz, Inc.). The sequencing results were used to determine the measles genotype as described in section 2.1.

### 2.4. Detection of measles, mumps, and rubella RNA in wastewater liquid and solid fractions

We compared the concentrations of the measles, mumps, and rubella RNA detected in the wastewater liquid and solid fractions by spiking Edmonston strain measles virus (VR-24, ATCC), mumps virus (VR-106, ATCC), and rubella virus (VR-1359, ATCC) into influent wastewater at a target concentration of 10^7^ copies/L-wastewater of each virus. The ATCC virus standards were first diluted and then spiked into 6 replicate wastewater samples. After spiking the virus standards, the wastewater samples were inverted to mix and sat at 4°C for 10 minutes, following a previously published procedure (Breadner et al. 2023). After, the wastewater samples were aliquoted into 50 mL tubes and centrifuged to separate liquid and solid fractions, as described in section 2.3. Six unspiked wastewater samples were processed in parallel as negative controls.

We calculated the partition coefficient (K_d_) of measles, mumps, and rubella viral RNA in influent samples as the ratio of the concentration of viral RNA in the solids (copies/g) divided by the concentration in the liquid (copies/mL) (SI. Eq 1). We also calculated the recovery rate of each target virus in the liquid and solid fraction of wastewater (SI. Eq 2). The Mann-Whitney U test was used to compare the log-transformed measles, mumps, and rubella RNA concentrations measured in liquid and solid fractions. In addition, we evaluated the presence of inhibitors in the sample fractions by diluting the RNA extracts 10-fold. We calculated an inhibition factor as the concentration of viral RNA based on 10x diluted extracts divided by the concentration of viral RNA from the undiluted extracts (SI. Eq 3).

### 2.5. Measles mumps, and rubella RNA persistence in wastewater

Persistence experiments were performed by spiking the Edmonston strain measles virus (VR-24, ATCC), mumps virus (VR-106, ATCC), and rubella virus (VR-1359, ATCC) into 400 mL of influent wastewater. Three replicate 400 mL-aliquots were placed in four different conditions for 28 days. We included two different target initial concentrations of each virus (“high” of approximately 10^7^ copies/L-wastewater, and “low” of 10^6^ copies/L-wastewater), and incubation at two temperatures (4°C and room temperature). Room temperature was not controlled but was recorded using a Traceable Temperature/Humidity Bluetooth Data Logger (6537CC, Traceable). Triplicate unspiked wastewater samples were also stored at the two temperatures as the negative controls. Samples were collected on days 0, 1, 3, 7, 14, 21, and 28 days for quantification of measles, mumps, and rubella RNA. On each measurement day, we used a serological pipette to gently mix each sample and collect 50 mL of wastewater from the mid-depth of each sample bottle. The liquid fraction of the samples were used for quantification and samples were processed as described in Section 2.2.

A first-order log-linear decay model was used to describe the persistence of each virus. Viral RNA concentrations were normalized against day 0 concentrations before fitting a linear regression. We excluded measurement results when no positive droplet was observed in two or more of the replicates. First-order decay rate constants (k) were estimated for each virus for each different condition (SI. Eq 4-5) and standard errors for each k based on the triplicates. We also calculated the T_90_ for each virus in each condition, which is the time required for the viral RNA concentration to be reduced by 1 log10 (90% reduction). A z-test was used to compare the first-order decay rates determined at the two different initial concentrations, using the mean and standard error calculated for each decay rate constant and assuming the rates are normally distributed ^49^.

## 3. Results and Discussion

### 3.1. Measles, mumps and rubella assay validation

We first validated the mumps, rubella, and measles WT1 multiplex assay using gblock synthetic gene fragments (Integrated DNA Technologies, Coralville, LA, USA). After confirming amplification and clear separation of gblock standards, we tested our assay on virus standards purchased from ATCC. These included the Edmonston strain measles virus (VR-24, ATCC), mumps virus (VR-106, ATCC), and rubella virus (VR-1359, ATCC) (Figures SI.1). We again observed clear separation of positive and negative droplets in standards and clean controls. We also verified that singleplex and multiplex RT-ddPCR assays yielded similar quantitative results and observed no significant difference between them using both gblocks and ATCC standards (p-values > 0.1; Figure SI.2 and Table SI.9). Finally, we tested the assays on wastewater samples spiked with the ATCC standards (Figure SI.1). For all the tests, we observed significant separations between the positive and negative droplets for all the targets based on the RT-ddPCR results, indicating that the multiplex assay can be used to detect measles, mumps, and rubella viruses in wastewater.

Next, to test whether our WT2 and VA assays could distinguish between wildtype and vaccine strains of measles, we performed experiments in which we spiked ATCC Edmonston vaccine and B3 measles virus strains individually and in a mixture into water and wastewater samples. The RT-ddPCR results showed that the amplitudes of the positive droplets could be used to distinguish between the detection of the wildtype B3 strain and the vaccine strain (Figure 1B and Table SI.10). Specifically, the amplitude of the wild-type strain was above 5,000 using the WT2 assay (channel 2) and below 5,000 when using the VA assay (channel 3), and the amplitude of the vaccine strain was above 6,000 when using the VA assay and below 3,000 when using the WT2 assay. When both strains were spiked into the wastewater, a clear separation was observed between the negative droplets, positive droplets corresponding to the wild-type strain, and positive droplets corresponding to the vaccine strain using both the WT2 and VA assays. More substantial separation between wild-type and vaccine-positive droplet clusters was observed with the WT2 assay than the VA assay.

**Figure 1.**
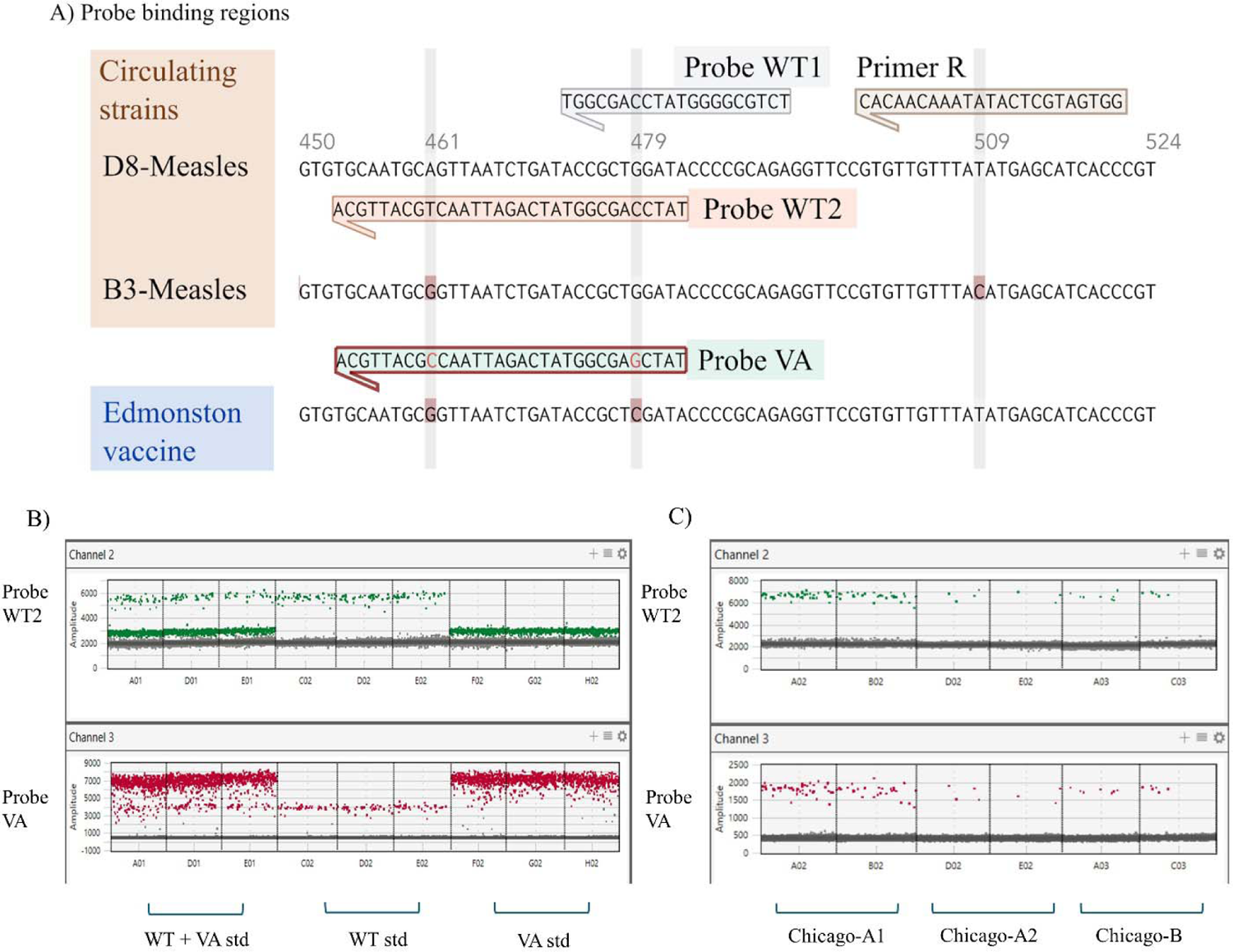
Measles assay development and validation. (A) Alignment of the M gene of measles virus genotypes D8, B3, and Edmonston vaccine (VA) strains. Assays for distinguishing between wild type (WT) strains (D8 and B3) and vaccine strains used the same forward and reverse primers and different probe sequences, shown above the aligned sequences. (B) Validation of the WT2 and vaccine assays using ATCC standards spiked into wastewater. ddPCR results for the WT2 probe (channel 2, top panel) showed a cluster of positive droplets between 5,000-6,000 amplitude, corresponding to presence of the WT standard (genotype B3), and a cluster with a 3,000 amplitude corresponding to the vaccine standard. The VA probe (channel 3, bottom panel) showed a positive cluster at 7,000 amplitude corresponding to the VA standard and a positive cluster at 4,000 corresponding to the WT standard. (C) Validation of the WT2 and vaccine assays using wastewater samples from a facility in Chicago, IL, with an active measles outbreak. All samples showed positive detections of WT strain, as indicated by the amplitude of the positive droplets in both the WT assay (channel 2, top panel) and VA assay (channel 3, bottom panel).

In addition to validating the assays with standards spiked into wastewater, we obtained a sample collected from a manhole located outside a facility with an ongoing measles outbreak. In these samples, the measles-positive droplets had an amplitude over 6,000 using the WT2 assay and an amplitude less than 2,500 using the VA assay, indicative that the sample contained the wild-type measles strain. We confirmed the detection and genotype of the wildtype strain by amplifying a region of the genome and performing Sanger sequencing. The Sanger sequencing results indicated that measles detected was most closely related to the D8 measles genotype (Figure SI.3), which is a dominant circulating strain in the current outbreak.

Here, we developed and validated a multiplex ddPCR assay that can detect measles, mumps, and rubella in wastewater samples. Further, our measles assays distinguish between the wild type and vaccine strains in wastewater. Using this assay, we were able to detect wild-type measles in wastewater samples containing endogenous measles RNA from a facility with an active outbreak (Figure 1C). Existing measles assays have been developed for wastewater monitoring ^40,50–52^. However, these assays are not able to distinguish between wild type and vaccine strains, which could potentially result in positive detections associated with vaccination events as opposed to transmission in communities. The current CDC recommended assays for the detection of measles in clinical samples use one assay to detect all measles strains (inclusive of wild type and vaccine strains) ^51^, and a second assay that targets a different region of the measles genome to detect vaccine strains only ^52^. Thus, to differentiate between a wild type and vaccine strain in a sample, both assays must be performed. Here, we present ddPCR assays that can distinguish between wild type and vaccine measles strains by amplitude differentiation. The WT2 and VAC assays use the same forward and reverse primers, but differ in probe sequence. The probes were designed to maximize binding affinity difference by placing them at mismatches between wild type and vaccine strains. This approach has been used widely in ddPCR assays for detecting single nucleotide polymorphisms (SNPs) in genes associated with cancer ^53,54^.

### 3.2. Evaluating recovery of measles, mumps, and rubella from liquid and solid fractions of influent wastewater samples

To understand how concentration and sample processing methods impacted the detection of measles, mumps, and rubella in wastewater samples, we quantified viral RNA concentrations in the liquid and solid fractions of wastewater samples spiked with virus standards. We observed significantly higher levels of all target RNA in the liquid than the solid fraction of wastewater (p-values < 0.01; Figure 2A). We also calculated the recovery rate for each target in each fraction of the wastewater (SI. Eq 2). The concentration of measles in the liquid fraction (supernatant) was 10 ^7.09 ± 0.18^ copies/L-wastewater, which corresponded to a recovery rate of 54.2 ± 13.4%. In contrast, the concentration of measles in the solid fraction (pellet) was 10 ^5.88 ± 0.24^ copies/L-wastewater, with a recovery rate of 3.7± 13.4%. Similar trends of higher concentrations in the liquid over the solid fraction were observed for mumps and rubella. For mumps, the concentration in the liquid fraction was 10 ^6.90^ ^± 0.17^ copies/L-wastewater (72.8 ± 24.9% recovery rate) and 10 ^5.07^ ^± 0.30^ copies/L-wastewater in the solids (1.20 ± 0.68% recovery rate). For rubella, the liquid concentration was 10 ^6.74^ ^± 0.21^ copies/L-wastewater (31.1 ± 12.3% recovery rate) and the solids concentration was 10 ^3.67^ ^± 0.37^ copies/L-wastewater (0.03 ± 0.02% recovery rate). Differences in recovery rates were in part due to differences in RT-ddPCR inhibition in liquid vs. solid extracts (Figure SI.4). When we diluted our nucleic acid extracts 10x, we observed minimal inhibitory effects in liquid (inhibition factors of 1.0 - 2.2) as compared to the solids sample extracts (inhibition factors of 5.7 - 32.4). However, even accounting for differences in inhibition, the liquid fraction yielded overall higher concentrations of all viruses as compared to the solid fraction.

**Figure 2.**
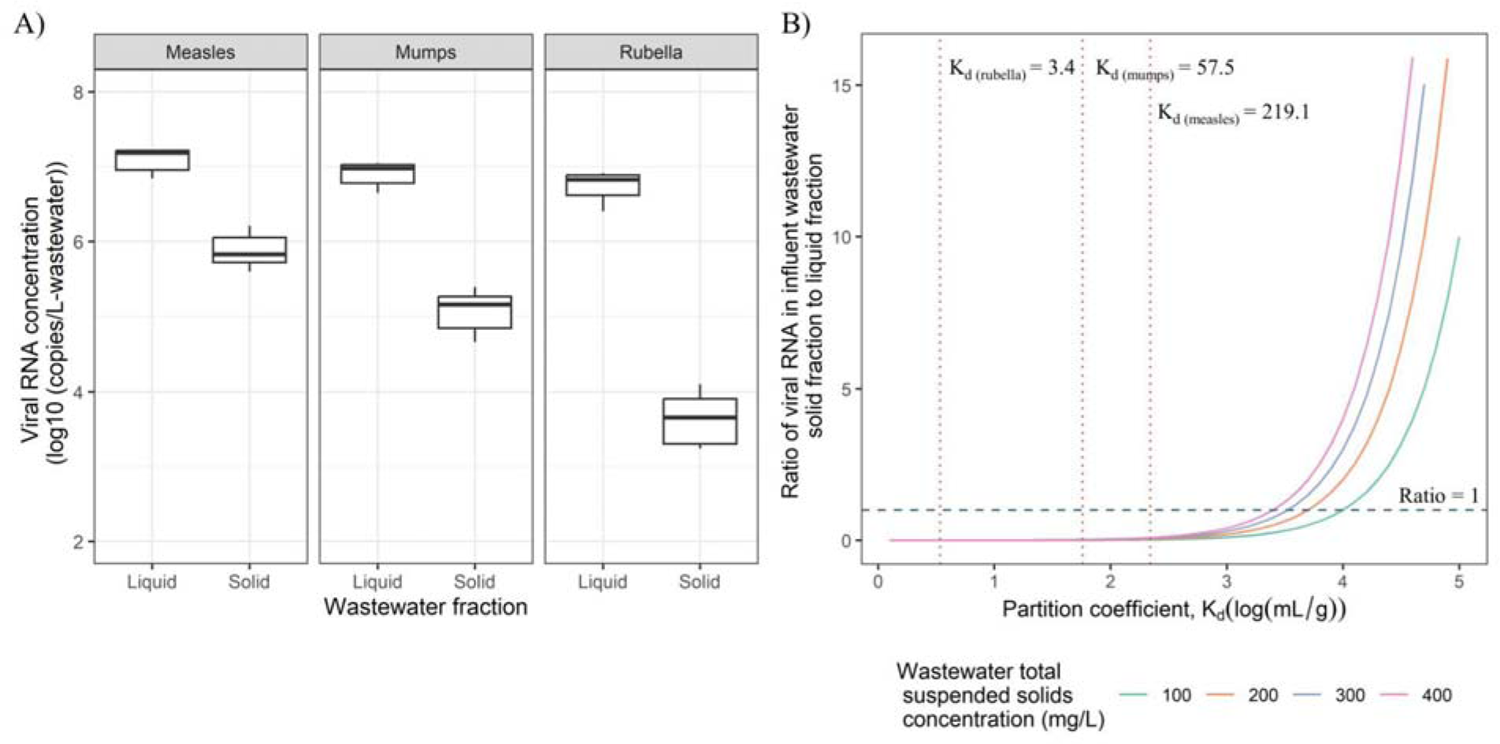
Partitioning of measles, mumps, and rubella viral RNA. (A) Measles, mumps, and rubella viral RNA concentrations in influent wastewater liquid and solid fractions. Boxplots show the results generated from 6 replicates of 50mL influent wastewater samples spiked with viral ATCC standards that were centrifuged and the supernatant (liquid) and pellets (solid) were processed separately for viral RNA quantification. (B) Impact of virus partition coefficient (K_d_) and total suspended solids concentration (TSS) in wastewater on relative amount of target viral RNA in solid vs. liquid fraction. The x-axis represents the log-transformed partition coefficient (mL/g) and the y-axis represents the ratio of the target viral RNA in the solid to the liquid fraction of a wastewater sample (viral RNA concentration (copies/L-wastewater) in solid fraction divided by viral RNA concentration (copies/L-wastewater) in liquid fraction). The blue dashed line indicates where the abundance of target virus in wastewater solid and liquid fractions are equal. A greater y-axis value indicates that the target virus is more abundant in wastewater solids than liquid fraction. Different colored lines represent different wastewater total suspended solids concentrations. Orange dotted lines indicate the K_d_ values of measles, mumps, and rubella determined in this study.

To assess the partitioning behavior of measles, mumps, and rubella viruses in the liquid vs. solid fractions of wastewater, we estimated the partition coefficients (K_d_) using samples spiked with viral standards. K_d_ was calculated as the ratio of *C*_s_/*C*_L_. We observed the highest K_d_ for measles of 219.1 ± 99.3 mL/g, as compared to 57.5 ± 38.6 mL/g for mumps and 3.4 ± 2.5 mL/g for rubella. These K_d_ results are lower than those determined in previous studies for other enveloped RNA viruses, such as SARS-CoV-2, influenza A, RSV, and Zika, in wastewater ^55^, ^56^. One possible reason is that using the PCR inhibitor removal kit minimized the inhibition during the partitioning experiment of Roldan-Hernandez et al. (2023, 2024) ^55^, ^56^, while significant inhibitory effects were observed in the solids sample extracts in our study. In addition, partition coefficients and recovery rates vary across different viruses, and also due to numerous other factors, such as the wastewater matrix, liquid-solid separation methods, and wastewater processing methods ^16,55–58^. Additional research is needed to investigate how these factors impact the partitioning and recovery of measles, mumps, and rubella in wastewater samples.

For all three viruses, viral RNA was enriched in the solids over the liquid fraction of wastewater on a per mass basis (i.e., gene copies/g of wastewater solids > gene copies/mL of wastewater liquid). Despite a higher mass of viral RNA in the solids, higher *concentrations* of viral RNA were measured in the liquid fraction of wastewater. This is because the concentration of RNA measured in the liquid and solid fraction of a wastewater sample depends on the solids concentration of the wastewater sample in addition to the virus partitioning behavior (Figure 2B). Concentrations of the viral RNA thus may be enriched in the solid fraction by orders of magnitude relative to the liquid phase and yet in a given volume of influent wastewater, most of the viruses are in the liquid phase. In the case of our influent wastewater samples, they contained a total suspended solids concentration of 316 mg/L-wastewater, and thus it makes sense that given the K_d_ values observed that we were able to more sensitively recover the target viral RNA in the liquid fraction of the wastewater (see dashed vertical lines on Figure 2B).

Inhibitors present in the sample may also drive decisions related to sample processing methods to improve sensitivity. As noted above, we observed significantly more inhibition in our solid fraction RNA extracts than liquid fraction extracts. Employing a concentration method that does not separate liquid and solid fractions, such as Ceres Nanotrap beads ^12,59–61^ or electronegative filtration of the entire sample without a prior centrifugation step ^24,62,63^, should be assessed to compare and balance the concentration of target viruses with inhibitors. RT-PCR inhibitor removal kits should also be explored for improving method sensitivity.

### 3.3. Measles, mumps, and rubella RNA persistence in wastewater

In the measles, mumps, and rubella RNA persistence experiments, we determined the viral RNA concentration of measles, mumps, and rubella at day 0 to be 0.25-1.66×10^7^ copies/L-wastewater for the high-concentration samples and 0.63-1.59×10^6^ copies/L-wastewater for the low-concentration samples (Figure 3). A first-order decay model was fit to the viral RNA concentration measurements over time for all viruses and for each different experimental condition (R^2^ values ranged between 0.72 and 0.98, p-values < 0.001). Details of the initial concentrations, the calculated first-order decay rate constants (k), T_90_, and the R^2^ values for each viral target and experimental condition are summarized in Table SI.11.

**Figure 3.**
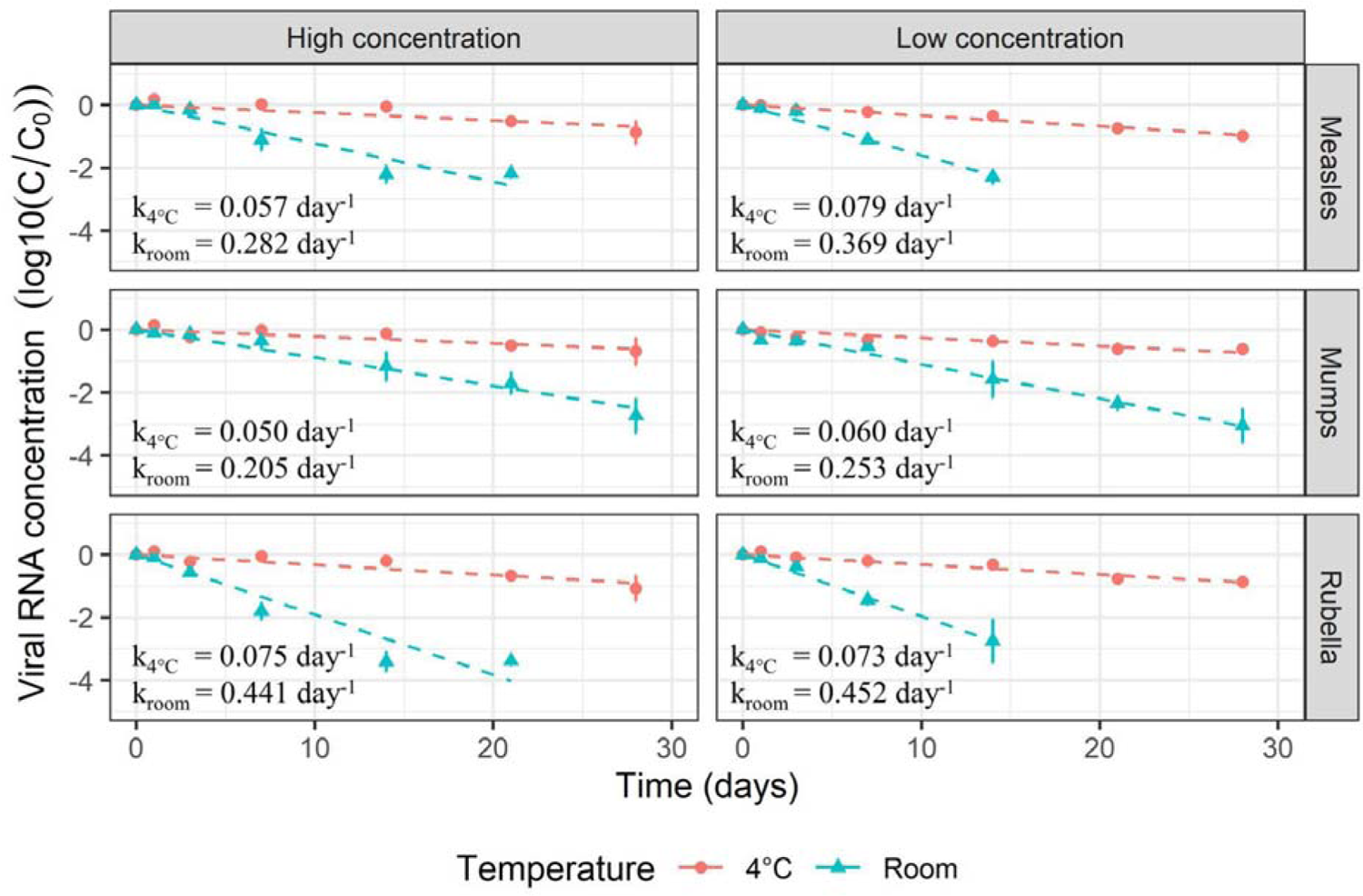
Measles, mumps, and rubella viral RNA persistence in influent wastewater over 28 days. The x-axis indicates the day of the persistence experiment. The y-axis indicates the normalized target RNA concentration (log10(C/C_0_), where C is the target RNA concentration on the measurement day and C_0_ is the concentration on day 0. Points and error bars indicate the mean values and standard deviations of the viral RNA concentrations from triplicate samples. Dashed lines show the fitted linear regression. First-order decay rate constants (k) are shown for each target viral RNA. Samples with no positive droplets in two or more of the triplicates were excluded.

When stored at 4°C, limited decay of measles, mumps, and rubella viral RNA was observed, with less than 90% reduction for all the target viruses and both initial concentrations over the 28-day test. The decay rate constants were between 0.050 ± 0.007 and 0.080 ± 0.013 day^-1^, and the time for one log reduction (T_90_) ranged from 29.3 to 45.7 days. The high-concentration mumps RNA had the lowest decay rate constant and the highest T_90_ value, while the low-concentration measles RNA had the highest decay rate constant and the lowest T_90_ value. The measles, mumps, and rubella viral RNA decayed faster in wastewater stored at room temperature, which ranged from 21°C to 26°C (Figure SI.5). Decay rate constants ranged from 0.205 ± 0.011 - 0.452 ± 0.024 day^-1^, with average T_90_ values of 5.1-11.2 days. The lowest decay rate constants were observed for mumps RNA, with mean values of 0.205 and 0.253 day^-1^, and the highest decay rate constants were observed for rubella RNA, with mean values of 0.441 and 0.452 day^-1^.

We observed significantly higher (faster) decay rate constants in the wastewater samples spiked with the low initial concentration of virus standards compared to the wastewater samples spiked with the high initial concentration of virus standards for measles at both 4°C and room temperature and for mumps at room temperature (p-values < 0.05). For measles RNA, the mean T_90_ value decreased from 40.5 to 29.3 days (27.7% decrease) when the samples were stored at 4°C and from 8.2 to 6.2 days (24.4% decrease) when the samples were stored at room temperature. For mumps RNA, the mean T_90_ values decreased from 11.2 to 9.1 days (18.8% decrease) when the samples were stored at room temperature. No significant differences between the two viral RNA levels were observed for rubella or when mumps were stored at 4°C. Similar results were also observed by Zhu et al. 2023 when they investigated the persistence of Zika spiked into wastewater at different initial concentrations, and additional studies are needed to further investigate viral RNA persistence at low viral concentrations ^49^.

To the best of our knowledge, this is the first study to report decay kinetics of measles, mumps, and rubella viral RNA in wastewater. Our results show that measles, mumps, and rubella RNA decay in wastewater is limited. Similar to other studies that investigated the persistence of enveloped RNA viruses in wastewater ^49,64–68^, we show that the viral RNA decayed significantly slower in samples stored at a lower temperature. These results also demonstrate the feasibility of wastewater monitoring of measles, mumps, and rubella viruses, and suggest limited decay of all viruses (T_90_ values were >5 days) at room temperature, indicating only minimal loss due to decay during wastewater transit within a building or facility and also within the sewer system. Our results also indicate that wastewater samples can be stored at 4°C for up to several weeks without substantial loss in viral RNA signal. Further studies are needed to investigate measles, mumps, and rubella RNA persistence during freeze-thaw cycles, at lower initial concentrations, at higher temperatures, and in different wastewaters that may vary in total suspended solids (TSS) concentrations, pH, and other water quality parameters.

Our study supports the use of wastewater monitoring for population-level surveillance of three vaccine-preventable diseases: measles, mumps, and rubella. We developed and validated multiplexed RT-ddPCR assays for the detection of wild type and vaccine measles strains, mumps, and rubella. We estimated partition coefficients for each virus in influent wastewater samples and showed that the viral RNA was enriched in the solid fraction relative to the liquid and yet given the low solids content, assaying the liquid fraction yielded more sensitive detection of the viruses. Finally, we measured the persistence of the viral RNA in wastewater samples at room temperature and 4°C and show that limited decay of the viral targets is expected over time scales relevant to sewage transit and storage. Together these findings suggest that wastewater monitoring for these vaccine-preventable diseases is feasible to integrate into existing routine wastewater surveillance systems. While cases of measles are still rare in the U.S., measles remains endemic in several other countries. With vaccination rates below those needed for herd immunity, cases of measles are on the rise and improved surveillance may be critical for early detection and assessing the extent and progression of an outbreak. Detections of these viruses can be used to dispatch and target immunization clinics and communication efforts to encourage vaccination.

We recommend several areas of further research on wastewater monitoring for measles mumps, and rubella viruses. First, we need validated assays that can distinguish between wild type and vaccine strains of mumps and rubella in addition to measles. We focused on measles due to the urgency from recent outbreaks. Second, studies on shedding of measles, mumps, and rubella viruses both during infection and after vaccination are needed to understand and model expected concentrations in wastewater and assess the sensitivity of wastewater monitoring for detecting cases in communities. Third, we need studies that compare monitoring at facilities to downstream wastewater treatment plants. Fourth, methods for improving the recovery of endogenous target viruses are needed. Fifth, studies on viral persistence as a function of different wastewater quality parameters and storage conditions (e.g., freeze/thaw) are needed to optimize sampling and laboratory methods. And finally, sequencing of wastewater viral targets should be explored for tracking disease transmission using genomic epidemiology.

## Data Availability

All data produced in the present study are available upon reasonable request to the authors and the sequencing data has been submitted to NCBI.

## Acknowledgments

This research was supported by funds from the Centers for Disease Control and Prevention (NI50CK000557) and the Houston Health Department. We thank Kaavya Domakonda, Rebecca Schneider, and Anthony Mulenga at the Houston Health Department and Houston Water for their assistance in sample collection. We also thank Dolores Sanchez Gonzalez and Michael Secreto for collecting and sending wastewater samples and an insightful conversation. We thank Lily Metsker for her help with literature review. The graphical abstract was created with Biorender.com.

## Author Contributions

**Jingjing Wu:** Data curation, Investigation, Visualization, Writing-original draft, Writing-review & editing

**Michael X. Wang:** Methodology, Visualization, Writing-original draft

**Prashant Kalvapalle:** Methodology, Visualization, Writing-original draft

**Michael Nute:** Methodology

**Todd Treangen:** Methodology, Funding acquisition, Supervision, Writing-review & editing

**Katherine Ensor:** Conceptualization, Funding acquisition, Supervision, Methodology, Writing-review & editing

**Loren Hopkins:** Conceptualization, Funding acquisition, Supervision, Writing-review & editing

**Rachel Poretsky:** Methodology, Writing-review & editing

**Lauren B. Stadler (Corresponding Author):** Conceptualization, Funding acquisition, Project administration, Supervision, Writing-original draft, Writing-review & editing

## References

1 P. Gahlot, K. D. Alley, S. Arora, S. Das, A. Nag and V. K. Tyagi, Wastewater surveillance could serve as a pandemic early warning system for COVID-19 and beyond, WIREs Water, 2023, 10, e1650.

2 B. Kasprzyk-Hordern, B. Adams, I. D. Adewale, F. O. Agunbiade, M. I. Akinyemi, E. Archer, F. A. Badru, J. Barnett, I. J. Bishop, M. Di Lorenzo, P. Estrela, J. Faraway, M. J. Fasona, S. A. Fayomi, E. J. Feil, L. J. Hyatt, A. T. Irewale, T. Kjeldsen, A. K. S. Lasisi, S. Loiselle, T. M. Louw, B. Metcalfe, S. A. Nmormah, T. O. Oluseyi, T. R. Smith, M. C. Snyman, T. O. Sogbanmu, D. Stanton-Fraser, S. Surujlal-Naicker, P. R. Wilson, G. Wolfaardt and C. O. Yinka-Banjo, Wastewater-based epidemiology in hazard forecasting and early-warning systems for global health risks, Environ. Int., 2022, 161, 107143.

3 M. Kumar, G. Jiang, A. Kumar Thakur, S. Chatterjee, T. Bhattacharya, S. Mohapatra, T. Chaminda, V. Kumar Tyagi, M. Vithanage, P. Bhattacharya, L. D. Nghiem, D. Sarkar, C. Sonne and J. Mahlknecht, Lead time of early warning by wastewater surveillance for COVID-19: Geographical variations and impacting factors, Chem. Eng. J., 2022, 441, 135936.

4 E. Róka, B. Khayer, Z. Kis, L. B. Kovács, E. Schuler, N. Magyar, T. Málnási, O. Oravecz, B. Pályi, T. Pándics and M. Vargha, Ahead of the second wave: Early warning for COVID-19 by wastewater surveillance in Hungary, Sci. Total Environ., 2021, 786, 147398.

5 M. Assoum, C. L. Lau, P. K. Thai, W. Ahmed, J. F. Mueller, K. V. Thomas, P. M. Choi, G. Jackson and L. A. Selvey, Wastewater Surveillance Can Function as an Early Warning System for COVID-19 in Low-Incidence Settings, Trop. Med. Infect. Dis., 2023, 8, 211.

6 D. Panchal, O. Prakash, P. Bobde and S. Pal, SARS-CoV-2: sewage surveillance as an early warning system and challenges in developing countries, Environ. Sci. Pollut. Res., 2021, 28, 22221–22240.

7 M. D. Parkins, B. E. Lee, N. Acosta, M. Bautista, C. R. J. Hubert, S. E. Hrudey, K. Frankowski and X.-L. Pang, Wastewater-based surveillance as a tool for public health action: SARS-CoV-2 and beyond, Clin. Microbiol. Rev., 2023, 0, e00103–22.

8 J. R. Thompson, Y. V. Nancharaiah, X. Gu, W. L. Lee, V. B. Rajal, M. B. Haines, R. Girones, L. C. Ng, E. J. Alm and S. Wuertz, Making waves: Wastewater surveillance of SARS-CoV-2 for population-based health management, Water Res., 2020, 184, 116181.

9 G. Medema, F. Been, L. Heijnen and S. Petterson, Implementation of environmental surveillance for SARS-CoV-2 virus to support public health decisions: Opportunities and challenges, Curr. Opin. Environ. Sci. Health, 2020, 17, 49–71.

10 J. Oghuan, C. Chavarria, S. R. Vanderwal, A. Gitter, A. A. Ojaruega, C. Monserrat, C. X. Bauer, E. L. Brown, S. J. Cregeen, J. Deegan, B. M. Hanson, M. Tisza, H. I. Ocaranza, J. Balliew, A. W. Maresso, J. Rios, E. Boerwinkle, K. D. Mena and F. Wu, 2023, 2023.05.28.23290658.

11 Y. Wang, P. Liu, J. VanTassell, S. P. Hilton, L. Guo, O. Sablon, M. Wolfe, L. Freeman, W. Rose, C. Holt, M. Browning, M. Bryan, L. Waller, P. F. M. Teunis and C. L. Moe, When case reporting becomes untenable: Can sewer networks tell us where COVID-19 transmission occurs?, Water Res., 2023, 229, 119516.

12 K. Brighton, S. Fisch, H. Wu, K. Vigil and T. G. Aw, Targeted community wastewater surveillance for SARS-CoV-2 and Mpox virus during a festival mass-gathering event, Sci. Total Environ., 2024, 906, 167443.

13 T. de Melo, G. Islam, D. B. D. Simmons, J.-P. Desaulniers and A. E. Kirkwood, An alternative method for monitoring and interpreting influenza A in communities using wastewater surveillance, Front. Public Health, 2023, 11, 1141136.

14 B. A. Layton, D. Kaya, C. Kelly, K. J. Williamson, D. Alegre, S. M. Bachhuber, P. G. Banwarth, J. W. Bethel, K. Carter, B. D. Dalziel, M. Dasenko, M. Geniza, A. George, A.-M. Girard, R. Haggerty, K. A. Higley, D. M. Hynes, J. Lubchenco, K. R. McLaughlin, F. J. Nieto, A. Noakes, M. Peterson, A. D. Piemonti, J. L. Sanders, B. M. Tyler and T. S. Radniecki, Evaluation of a Wastewater-Based Epidemiological Approach to Estimate the Prevalence of SARS-CoV-2 Infections and the Detection of Viral Variants in Disparate Oregon Communities at City and Neighborhood Scales, Environ. Health Perspect., 2022, 130, 067010.

15 S. Karthikeyan, N. Ronquillo, P. Belda-Ferre, D. Alvarado, T. Javidi, C. A. Longhurst and R. Knight, High-Throughput Wastewater SARS-CoV-2 Detection Enables Forecasting of Community Infection Dynamics in San Diego County, mSystems, 2021, 6, 10.1128/msystems.00045-21.

16 E. Mercier, P. M. D’Aoust, O. Thakali, N. Hegazy, J.-J. Jia, Z. Zhang, W. Eid, J. Plaza-Diaz, M. P. Kabir, W. Fang, A. Cowan, S. E. Stephenson, L. Pisharody, A. E. MacKenzie, T. E. Graber, S. Wan and R. Delatolla, Municipal and neighbourhood level wastewater surveillance and subtyping of an influenza virus outbreak, Sci. Rep., 2022, 12, 15777.

17 P. M. D’Aoust, E. Mercier, D. Montpetit, J.-J. Jia, I. Alexandrov, N. Neault, A. T. Baig, J. Mayne, X. Zhang, T. Alain, M.-A. Langlois, M. R. Servos, M. MacKenzie, D. Figeys, A. E. MacKenzie, T. E. Graber and R. Delatolla, Quantitative analysis of SARS-CoV-2 RNA from wastewater solids in communities with low COVID-19 incidence and prevalence, Water Res., 2021, 188, 116560.

18 B. Hughes, D. Duong, B. J. White, K. R. Wigginton, E. M. G. Chan, M. K. Wolfe and A. B. Boehm, Respiratory Syncytial Virus (RSV) RNA in Wastewater Settled Solids Reflects RSV Clinical Positivity Rates, Environ. Sci. Technol. Lett., 2022, 9, 173–178.

19 Y. Ai, A. Davis, D. Jones, S. Lemeshow, H. Tu, F. He, P. Ru, X. Pan, Z. Bohrerova and J. Lee, Wastewater SARS-CoV-2 monitoring as a community-level COVID-19 trend tracker and variants in Ohio, United States, Sci. Total Environ., 2021, 801, 149757.

20 B. Kasprzyk-Hordern, N. Sims, K. Farkas, K. Jagadeesan, K. Proctor, M. J. Wade and D. L. Jones, Wastewater-based epidemiology for comprehensive community health diagnostics in a national surveillance study: Mining biochemical markers in wastewater, J. Hazard. Mater., 2023, 450, 130989.

21 M. K. Wolfe, A. T. Yu, D. Duong, M. S. Rane, B. Hughes, V. Chan-Herur, M. Donnelly, S. Chai, B. J. White, D. J. Vugia and A. B. Boehm, Use of Wastewater for Mpox Outbreak Surveillance in California, N. Engl. J. Med., 2023, 388, 570–572.

22 J. Crowe, A. T. Schnaubelt, S. SchmidtBonne, K. Angell, J. Bai, T. Eske, M. Nicklin, C. Pratt, B. White, B. Crotts-Hannibal, N. Staffend, V. Herrera, J. Cobb, J. Conner, J. Carstens, J. Tempero, L. Bouda, M. Ray, J. V. Lawler, W. S. Campbell, J.-M. Lowe, J. Santarpia, S. Bartelt-Hunt, M. Wiley, D. Brett-Major, C. Logan and M. J. Broadhurst, Assessment of a Program for SARS-CoV-2 Screening and Environmental Monitoring in an Urban Public School District, JAMA Netw. Open, 2021, 4, e2126447.

23 R. Fielding-Miller, S. Karthikeyan, T. Gaines, R. S. Garfein, R. A. Salido, V. J. Cantu, L. Kohn, N. K. Martin, A. Wynn, C. Wijaya, M. Flores, V. Omaleki, A. Majnoonian, P. Gonzalez-Zuniga, M. Nguyen, A. V. Vo, T. Le, D. Duong, A. Hassani, S. Tweeten, K. Jepsen, B. Henson, A. Hakim, A. Birmingham, P. D. Hoff, A. M. Mark, C. A. Nasamran, S. B. Rosenthal, N. Moshiri, K. M. Fisch, G. Humphrey, S. Farmer, H. M. Tubb, T. Valles, J. Morris, J. Kang, B. Khaleghi, C. Young, A. D. Akel, S. Eilert, J. Eno, K. Curewitz, L. C. Laurent, T. Rosing, R. Knight, N. A. Baer, T. Barber, A. Castro-Martinez, M. Chacón, W. Cheung, E. S. Crescini, E. R. Eisner, L. F. Vargas, A. Hakim, C. Hobbs, A. L. Lastrella, E. S. Lawrence, N. L. Matteson, K. Gangavarapu, T. T. Ngo, P. Seaver, E. W. Smoot, R. Tsai, B. Xia, S. Aigner, C. Anderson, P. Belda-Ferre, S. Sathe, M. Zeller, K. G. Andersen, G. W. Yeo and E. Kurzban, Safer at school early alert: an observational study of wastewater and surface monitoring to detect COVID-19 in elementary schools, Lancet Reg. Health – Am., DOI:10.1016/j.lana.2023.100449.

24 C. Gibas, K. Lambirth, N. Mittal, M. A. I. Juel, V. B. Barua, L. Roppolo Brazell, K. Hinton, J. Lontai, N. Stark, I. Young, C. Quach, M. Russ, J. Kauer, B. Nicolosi, D. Chen, S. Akella, W. Tang, J. Schlueter and M. Munir, Implementing building-level SARS-CoV-2 wastewater surveillance on a university campus, Sci. Total Environ., 2021, 782, 146749.

25 S. Kennedy and A. Spaulding, Four Models of Wastewater-Based Surveillance for SARS-CoV-2 in Jail Settings: How Monitoring Wastewater Complements Individual Screening, medRxiv, 2023, 2023.08.04.23293152.

26 B. A. Petros, J. S. Paull, C. H. Tomkins-Tinch, B. C. Loftness, K. C. DeRuff, P. Nair, G. L. Gionet, A. Benz, T. Brock-Fisher, M. Hughes, L. Yurkovetskiy, S. Mulaudzi, E. Leenerman, T. Nyalile, G. K. Moreno, I. Specht, K. Sani, G. Adams, S. V. Babet, E. Baron, J. T. Blank, C. Boehm, Y. Botti-Lodovico, J. Brown, A. R. Buisker, T. Burcham, L. Chylek, P. Cronan, A. Dauphin, V. Desreumaux, M. Doss, B. Flynn, A. Gladden-Young, O. Glennon, H. D. Harmon, T. V. Hook, A. Kary, C. King, C. Loreth, L. Marrs, K. J. McQuade, T. T. Milton, J. M. Mulford, K. Oba, L. Pearlman, M. Schifferli, M. J. Schmidt, G. M. Tandus, A. Tyler, M. E. Vodzak, K. K. Bevill, A. Colubri, B. L. MacInnis, A. Z. Ozsoy, E. Parrie, K. Sholtes, K. J. Siddle, B. Fry, J. Luban, D. J. Park, J. Marshall, A. Bronson, S. F. Schaffner and P. C. Sabeti, Multimodal surveillance of SARS-CoV-2 at a university enables development of a robust outbreak response framework, Med, 2022, 3, 883–900.e13.

27 L. C. Scott, A. Aubee, L. Babahaji, K. Vigil, S. Tims and T. G. Aw, Targeted wastewater surveillance of SARS-CoV-2 on a university campus for COVID-19 outbreak detection and mitigation, Environ. Res., 2021, 200, 111374.

28 R. R. Spurbeck, A. Minard-Smith and L. Catlin, Feasibility of neighborhood and building scale wastewater-based genomic epidemiology for pathogen surveillance, Sci. Total Environ., 2021, 789, 147829.

29 V. Vo, A. Harrington, C.-L. Chang, H. Baker, M. A. Moshi, N. Ghani, J. Y. Itorralba, R. L. Tillett, E. Dahlmann, N. Basazinew, R. Gu, T. D. Familara, S. Boss, F. Vanderford, M. Ghani, A. J. Tang, A. Matthews, K. Papp, E. Khan, C. Koutras, H.-Y. Kan, C. Lockett, D. Gerrity and E. C. Oh, Identification and genome sequencing of an influenza H3N2 variant in wastewater from elementary schools during a surge of influenza A cases in Las Vegas, Nevada, Sci. Total Environ., 2023, 872, 162058.

30 M. K. Wolfe, D. Duong, K. M. Bakker, M. Ammerman, L. Mortenson, B. Hughes, P. Arts, A. S. Lauring, W. J. Fitzsimmons, E. Bendall, C. E. Hwang, E. T. Martin, B. J. White, A. B. Boehm and K. R. Wigginton, Wastewater-Based Detection of Two Influenza Outbreaks, Environ. Sci. Technol. Lett., 2022, 9, 687–692.

31 M. Wolken, T. Sun, C. McCall, R. Schneider, K. Caton, C. Hundley, L. Hopkins, K. Ensor, K. Domakonda, P. Kalvapalle, D. Persse, S. Williams and L. B. Stadler, Wastewater surveillance of SARS-CoV-2 and influenza in preK-12 schools shows school, community, and citywide infections, Water Res., 2023, 231, 119648.

32 WHO, Measles, https://www.who.int/news-room/fact-sheets/detail/measles, (accessed 17 May 2024).

33 D. Kanbayashi, T. Kurata, A. Kaida, H. Kubo, S. P. Yamamoto, K. Egawa, Y. Hirai, K. Okada, Y. Kaida, R. Ikemori, T. Yumisashi, A. Ito, T. Saito, Y. Yamaji, Y. Nishino, R. Omori, H. Mori, K. Motomura and K. Ikuta, Shedding of rubella virus in postsymptomatic individuals; viral RNA load is a potential indicator to estimate candidate patients excreting infectious rubella virus, J. Clin. Virol., 2023, 160, 105377.

34 S. Gouma, S. J. M. Hahné, D. B. Gijselaar, M. P. G. Koopmans and R. S. van Binnendijk, Severity of mumps disease is related to MMR vaccination status and viral shedding, Vaccine, 2016, 34, 1868–1873.

35 R. S. van Binnendijk, S. van den Hof, H. van den Kerkhof, R. H. G. Kohl, F. Woonink, G. A. M. Berbers, M. A. E. Conyn-van Spaendonck and T. G. Kimman, Evaluation of Serological and Virological Tests in the Diagnosis of Clinical and Subclinical Measles Virus Infections during an Outbreak of Measles in The Netherlands, J. Infect. Dis., 2003, 188, 898–903.

36 M. J. Broadhurst, N. Garamani, Z. Hahn, B. Jiang, J. Weber, C. Huang, M. K. Sahoo, J. Kurzer, C. A. Hogan and B. A. Pinsky, Evaluation of a measles virus multiplex, triple-target real-time RT-PCR in three specimen matrices at a U.S. academic medical center, J. Clin. Virol., 2021, 136, 104757.

37 K. S. M. Benschop, H. G. van der Avoort, E. Jusic, H. Vennema, R. van Binnendijk and E. Duizer, Polio and Measles Down the Drain: Environmental Enterovirus Surveillance in the Netherlands, 2005 to 2015, Appl. Environ. Microbiol., 2017, 83, e00558–17.

38 K. Bibby and J. Peccia, Identification of viral pathogen diversity in sewage sludge by metagenome analysis, Environ. Sci. Technol., 2013, 47, 1945–1951.

39 C. McCall, H. Wu, B. Miyani and I. Xagoraraki, Identification of multiple potential viral diseases in a large urban center using wastewater surveillance, Water Res., 2020, 184, 116160.

40 A. Rector, M. Bloemen, B. Hoorelbeke, M. V. Ranst and E. Wollants, 2024, 2024.04.08.24305478.

41 F. Armas, F. Chandra, W. L. Lee, X. Gu, H. Chen, A. Xiao, M. Leifels, S. Wuertz, E. J. Alm and J. Thompson, Contextualizing Wastewater-Based surveillance in the COVID-19 vaccination era, Environ. Int., 2023, 171, 107718.

42 B. Kaic, I. Gjenero-Margan, B. Aleraj, T. Vilibić-Čavlek, M. Santak, A. Cvitković, T. Nemeth-Blazic and I. I. Hofman, Spotlight on measles 2010: Excretion of vaccine strain measles virus in urine and pharyngeal secretions of a child with vaccine associated febrile rash illness, Croatia, March 2010, Eurosurveillance, 2010, 15, 19652.

43 P. A. Rota, A. S. Khan, E. Durigon, T. Yuran, Y. S. Villamarzo and W. J. Bellini, Detection of measles virus RNA in urine specimens from vaccine recipients, J. Clin. Microbiol., 1995, 33, 2485–2488.

44 C. Camacho, G. Coulouris, V. Avagyan, N. Ma, J. Papadopoulos, K. Bealer and T. L. Madden, BLAST+: architecture and applications, BMC Bioinformatics, 2009, 10, 421.

45 K. Katoh, J. Rozewicki and K. D. Yamada, MAFFT online service: multiple sequence alignment, interactive sequence choice and visualization, Brief. Bioinform., 2019, 20, 1160–1166.

46 I. Letunic and P. Bork, Interactive Tree of Life (iTOL) v6: recent updates to the phylogenetic tree display and annotation tool, Nucleic Acids Res., 2024, gkae268.

47 E. G. Lou, N. Sapoval, C. McCall, L. Bauhs, R. Carlson-Stadler, P. Kalvapalle, Y. Lai, K. Palmer, R. Penn, W. Rich, M. Wolken, P. Brown, K. B. Ensor, L. Hopkins, T. J. Treangen and L. B. Stadler, Direct comparison of RT-ddPCR and targeted amplicon sequencing for SARS-CoV-2 mutation monitoring in wastewater, Sci. Total Environ., 2022, 833, 155059.

48 M. A. Borchardt, A. B. Boehm, M. Salit, S. K. Spencer, K. R. Wigginton and R. T. Noble, The Environmental Microbiology Minimum Information (EMMI) Guidelines: qPCR and dPCR Quality and Reporting for Environmental Microbiology, Environ. Sci. Technol., 2021, 55, 10210–10223.

49 K. Zhu, C. Hill, A. Muirhead, M. Basu, J. Brown, M. A. Brinton, M. J. Hayat, C. Venegas-Vargas, M. G. Reis, A. Casanovas-Massana, J. S. Meschke, A. I. Ko, F. Costa and C. E. Stauber, Zika virus RNA persistence and recovery in water and wastewater: An approach for Zika virus surveillance in resource-constrained settings, Water Res., 2023, 241, 120116.

50 E. K. Hayes, M. T. Gouthro, J. J. LeBlanc and G. A. Gagnon, Simultaneous detection of SARS-CoV-2, influenza A, respiratory syncytial virus, and measles in wastewater by multiplex RT-qPCR, Sci. Total Environ., 2023, 889, 164261.

51 K. B. Hummel, L. Lowe, W. J. Bellini and P. A. Rota, Development of quantitative gene-specific real-time RT-PCR assays for the detection of measles virus in clinical specimens, J. Virol. Methods, 2006, 132, 166–173.

52 F. Roy, L. Mendoza, J. Hiebert, R. J. McNall, B. Bankamp, S. Connolly, A. Lüdde, N. Friedrich, A. Mankertz, P. A. Rota and A. Severini, Rapid Identification of Measles Virus Vaccine Genotype by Real-Time PCR, J. Clin. Microbiol., 2017, 55, 735–743.

53 V. Rowlands, A. J. Rutkowski, E. Meuser, T. H. Carr, E. A. Harrington and J. C. Barrett, Optimisation of robust singleplex and multiplex droplet digital PCR assays for high confidence mutation detection in circulating tumour DNA, Sci. Rep., 2019, 9, 12620.

54 D. Schleinitz, J. K. DiStefano and P. Kovacs, in Disease Gene Identification: Methods and Protocols, ed. J. K. DiStefano, Humana Press, Totowa, NJ, 2011, pp. 77–87.

55 L. Roldan-Hernandez, C. V. Oost and A. B. Boehm, Solid–liquid partitioning of dengue, West Nile, Zika, hepatitis A, influenza A, and SARS-CoV-2 viruses in wastewater from across the USA, Environ. Sci. Water Res. Technol., DOI:10.1039/D4EW00225C.

56 L. Roldan-Hernandez and A. B. Boehm, Adsorption of Respiratory Syncytial Virus, Rhinovirus, SARS-CoV-2, and F+ Bacteriophage MS2 RNA onto Wastewater Solids from Raw Wastewater, Environ. Sci. Technol., 2023, 57, 13346–13355.

57 P. R. Breadner, H. A. Dhiyebi, A. Fattahi, N. Srikanthan, S. Hayat, M. G. Aucoin, S. J. Boegel, L. M. Bragg, P. M. Craig, Y. Xie, J. P. Giesy and M. R. Servos, A comparative analysis of the partitioning behaviour of SARS-CoV-2 RNA in liquid and solid fractions of wastewater, Sci. Total Environ., 2023, 895, 165095.

58 M. F. Espinosa, M. E. Verbyla, L. Vassalle, C. Leal, D. Leroy-Freitas, E. Machado, L. Fernandes, A. T. Rosa-Machado, J. Calábria, C. Chernicharo and C. R. Mota Filho, Reduction and liquid-solid partitioning of SARS-CoV-2 and adenovirus throughout the different stages of a pilot-scale wastewater treatment plant, Water Res., 2022, 212, 118069.

59 W. Ahmed, A. Bivins, A. Korajkic, S. Metcalfe, W. J. M. Smith and S. L. Simpson, Comparative analysis of Adsorption-Extraction (AE) and Nanotrap® Magnetic Virus Particles (NMVP) workflows for the recovery of endogenous enveloped and non-enveloped viruses in wastewater, Sci. Total Environ., 2023, 859, 160072.

60 M. Jiang, A. L. W. Wang, N. A. Be, N. Mulakken, K. L. Nelson and R. S. Kantor, Evaluation of the Impact of Concentration and Extraction Methods on the Targeted Sequencing of Human Viruses from Wastewater, Environ. Sci. Technol., 2024, 58, 8239–8250.

61 P. Liu, L. Guo, M. Cavallo, C. Cantrell, S. P. Hilton, A. Nguyen, A. Long, J. Dunbar, R. Barbero, R. Barclay, O. Sablon, M. Wolfe, B. Lepene and C. Moe, Comparison of Nanotrap® Microbiome A Particles, membrane filtration, and skim milk workflows for SARS-CoV-2 concentration in wastewater, Front. Microbiol., DOI:10.3389/fmicb.2023.1215311.

62 W. Ahmed, P. M. Bertsch, A. Bivins, K. Bibby, K. Farkas, A. Gathercole, E. Haramoto, P. Gyawali, A. Korajkic, B. R. McMinn, J. F. Mueller, S. L. Simpson, W. J. M. Smith, E. M. Symonds, K. V. Thomas, R. Verhagen and M. Kitajima, Comparison of virus concentration methods for the RT-qPCR-based recovery of murine hepatitis virus, a surrogate for SARS-CoV-2 from untreated wastewater, Sci. Total Environ., 2020, 739, 139960.

63 M. Ciesielski, D. Blackwood, T. Clerkin, R. Gonzalez, H. Thompson, A. Larson and R. Noble, Assessing sensitivity and reproducibility of RT-ddPCR and RT-qPCR for the quantification of SARS-CoV-2 in wastewater, J. Virol. Methods, 2021, 297, 114230.

64 W. Ahmed, P. M. Bertsch, K. Bibby, E. Haramoto, J. Hewitt, F. Huygens, P. Gyawali, A. Korajkic, S. Riddell, S. P. Sherchan, S. L. Simpson, K. Sirikanchana, E. M. Symonds, R. Verhagen, S. S. Vasan, M. Kitajima and A. Bivins, Decay of SARS-CoV-2 and surrogate murine hepatitis virus RNA in untreated wastewater to inform application in wastewater-based epidemiology, Environ. Res., 2020, 191, 110092.

65 F. Chandra, W. L. Lee, F. Armas, M. Leifels, X. Gu, H. Chen, S. Wuertz, E. J. Alm and J. Thompson, Persistence of Dengue (Serotypes 2 and 3), Zika, Yellow Fever, and Murine Hepatitis Virus RNA in Untreated Wastewater, Environ. Sci. Technol. Lett., 2021, 8, 785– 791.

66 A.-M. Hokajärvi, A. Rytkönen, A. Tiwari, A. Kauppinen, S. Oikarinen, K.-M. Lehto, A. Kankaanpää, T. Gunnar, H. Al-Hello, S. Blomqvist, I. T. Miettinen, C. Savolainen-Kopra and T. Pitkänen, The detection and stability of the SARS-CoV-2 RNA biomarkers in wastewater influent in Helsinki, Finland, Sci. Total Environ., 2021, 770, 145274.

67 A. Muirhead, K. Zhu, J. Brown, M. Basu, M. A. Brinton, F. Costa, M. J. Hayat and C. E. Stauber, Zika Virus RNA Persistence in Sewage, Environ. Sci. Technol. Lett., 2020, 7, 659–664.

68 A. I. Silverman and A. B. Boehm, Systematic Review and Meta-Analysis of the Persistence of Enveloped Viruses in Environmental Waters and Wastewater in the Absence of Disinfectants, Environ. Sci. Technol., 2021, 55, 14480–14493.

