## Supplementary Information for "Multiplexed detection, partitioning, and persistence of wild type and vaccine strains of measles, mumps, and rubella viruses in wastewater"

### SI 1. Materials and Methods

#### SI 1.1 Assay development for measles, mumps and rubella viruses

For measles, 105 complete D8 genomes (Supplementary Data 1) and 55 complete B3 genomes (Supplementary Data 2) were downloaded from the NCBI Virus database [1], with sample collection dates between Jan. 1, 2019 to Mar. 10, 2024, as well as 7 vaccine genomes (NCBI accessions: AF266286, AF266287, AF266288, AF266289, AF266291, FJ416067, DQ345723). A multiple sequence alignment (MSA) of all genomes above was generated with MAFFT [2] using default parameters. The primer pair and the WT1 probe were designed within a conserved region of the M gene with Primer3Plus [3]. WT2 and VA probes were designed within a region that was conserved among the wild type genomes yet bearing mismatches among the vaccine genomes.

For rubella, 63 complete genomes (Supplementary Data 3) were downloaded from the NCBI Virus database, with sample collection dates between Jan. 1, 2010 to Apr. 18, 2023. MSA was generated and the primer pair and the probe were designed within a conserved region of the start of RUBVgp1 gene with Primer3Plus.

For mumps, 108 complete genomes (Supplementary Data 4) were downloaded from the NCBI Virus database, with sample collection dates between Jan. 1, 2018 to Jun. 6, 2023. MSA was generated and the primer pair and the probe were designed within a conserved region of the L gene with Primer3Plus. The probe was manually redesigned to avoid starting with guanine.

Primer3Plus settings were as follows: product size of 70-150bp, primer size of 18-30bp (optimal 23bp), primer Tm (melting temperature) of 53-57°C (optimal 55°C), probe Tm (melting temperature) of 59-63°C (optimal 61°C), with all other parameters set to default. Output primers and probes were validated with NCBI BLAST+ [4] against the nucleotide collection (nt/nr) database, with the target species excluded, to ensure no non-specific amplification.

For the Sanger sequencing primers of measles, a conserved region of the L gene was input to Primer3Plus with settings: product size of 400-500bp, primer size of 18-35bp (optimal 25bp), primer Tm (melting temperature) of 58-62°C (optimal 60°C).

**SI 1.2 WWTP influent liquid and solid fraction concentration and target viral RNA quantification**

Wastewater samples were homogenized by inverting the bottle containing the sample several times and then aliquoting the sample into 50 mL centrifuge tubes. Tubes containing wastewater were centrifuged at 4,100 g and 4ºC for 20 minutes to separate liquid and solid fractions. The wastewater liquid fraction concentration was described previously by Laturner et al. (2021) and Lou et al. (2022)[5,6] and summarized as follows. After centrifugation, the supernatant was carefully poured into an MF 3, 300ml Magnetic Filter Holder with lid kit (20030001, Sterlitech) attached to the Multi-Vac 600-MS Manifold (180600-01, Sterlitech) and Rocker 800 Oil Free Laboratory Vacuum Pump (167800, Sterlitech) system without disturbing the pellet. The Electronegative Microbiological Analysis Membrane HA Filter (HAWG047S6, Millipore Sigma) was placed into the manifold system prior to the sample addition. One mL of 1.25 M $MgCl_2 \cdot H_2O$ (M0250, Sigma Aldrich) solution was added to the sample in the filter cup and then gently swirled with the pipette tip to mix and allowed to stand for 5 minutes before the vacuum pump was turned on. After all the sample passed through the filter, the filter was folded and transferred into a filled bead beating tube containing 0.1 mm diameter glass beads and 1 mL of lysis buffer from the chemagic™ Prime Viral DNA/RNA 300 Kit H96 (CMG-1433,

PerkinElmer), which was used for nucleic acid extraction. For the wastewater solid fraction, the pellets that remained at the bottom of the tube were resuspended using 1 mL of lysis buffer from the chemagic™ Prime Viral DNA/RNA 300 Kit H96 (CMG-1433, PerkinElmer) and transferred into a filled bead beating tube containing 0.1 mm diameter glass beads.

Bead beating tubes containing filters or pellets were beaten for 1 minute at 3,500 oscillations/m, two times using the Mini-Beadbeater 24 (112011, BioSpec) with a 2-minute break, where the tubes were placed on ice. After bead beating, the tubes were centrifuged at 17,000 g and 4°C for 5 minutes. 300 µL of lysate was used as input to nucleic acids extraction using Chemagic™ Prime Viral DNA/RNA 300 Kit H96 (Chemagic, CMG-1433, PerkinElmer), following the manufacturer's protocol, and the extracted nucleic acids were eluted in 50 µL of sterile, nuclease-free water. The extracts were stored at 4°C for no more than 2 hours before quantification.

The concentrations of target viral RNA were quantified using one-step RT-ddPCR Advanced Kit for Probes (1864021, Bio-Rad). Droplet generation was performed using an Automated Droplet Generator (1864101, Bio-Rad) and RT-ddPCR was completed using a C1000 ThermalCycler (Bio-Rad) and QX600 AutoDG Droplet Digital PCR System (Bio-Rad). Results were analyzed using QuantaSoft v1.7.4 software.

#### SI 1.3 RT-ddPCR assays setup and thermal cycling conditions

**Table SI.1:** Primers, probes, and standards used for quantification of measles, mumps, and rubella with RT-ddPCR

| **Target** | **Assay Name** | **Sequence (5'-3') / Catalogue #** | **Accession (coordinates)** |
|---|---|---|---|
| Measles (M gene) | Forward Primer | AATGAAAAACTGGTGTTCTACAA | NC_001498.1 (3759..3781) |

| | | | |
|---|---|---|---|
| | Reverse Primer | GGTGATGCTCATATAAACAACAC | NC_001498.1 (3926..3904) |
| | Probe – WT1 | /5SUN/TCTGCGGGG/ZEN/TATCCAGCGGT/3IABkFQ/ | NC_001498.1 (3897..3878) |
| | Probe – WT2 | /5SUN/TATCCAGCG/ZEN/GTATCAGATTAACTGCATTGCA/3IABkFQ/ | NC_001498.1 (3888..3858) |
| | Probe – VA | /5Cy5/TATCGAGCG/TAO/GTATCAGATTAACCGCATTGCA/3IAbRQSp/ | NC_001498.1 (3888..3858) |
| | Amplicon length | 168 | |
| | Standard (Vaccine) | VR-24 (ATCC) | |
| | Standard (Wildtype) | VR-1981(ATCC) | |
| Mumps (L gene) | Forward Primer | CATCTGGGCCAATTATAACTAC | NC_002200.1 (13312..13333) |
| | Reverse Primer | GATGTTTGTCTTTTCAATCTGAA | NC_002200.1 (13444..13422) |
| | Probe | /56-FAM/TATGTCACC/ZEN/TGAGGACAAATGTCAG/3IABkFQ/ | NC_002200.1 (13351..13375) |
| | Amplicon length | 133 | |
| | Standard | VR-106 (ATCC) | |
| Rubella (RUBVgp1 gene) | Forward Primer | CAGATGCAGGTTAGTGATCA | NC_076948.1 (218..237) |
| | Reverse Primer | GACGTGTAGGGCTTCTTTAG | NC_076948.1 (313..294) |
| | Probe | /5Cy5/CCCGCCGCC/TAO/ATTGGATCGAG/3IAbRQSp/ | NC_076948.1 (267..286) |
| | Amplicon length | 96 | |
| | Standard | VR-1359 (ATCC) | |
| Measles (M gene), mumps (L gene), and rubella (RUBVgp1 gene) | Gblock sequence | AATGAAAAACTGGTGTTCTACAACAACACCCCACTAACTCTCCTCACACCTTGGAGAAAGGTCCTAACAACAGGGAGTGTCTTCAACGCAAACCAAGTGTGCAATGCAGTTAATCTGATACCGCTGGATACCCCGCAGAGGTTCCGTGTTGTTTATATGAGCATCACCTCATCTGGGCCAATTATAACTACCCTCCAAAGGTTAAAGGTATGTCACCTGAGGACAAATGTCAGGCTTTAACTACCCATCTACTCCAAACTGTCGAATATGTTGAGCACATTCAGATTGAAAAGACAAACATCCAGATGCAGGTTAGTGATCACCCAGCACTCCAC | |

| | | |
|---|---|---|
| | | GCAATTTCGCGGTATACCCGCCGCCA TTGGATCGAGTGGGGCCCTAAAGAA GCCCTACACGTC |

**Table SI.2:** Final concentrations of the primer-probe mix for each target

| Assay Name | Final concentration (μM) | 40x concentration (μM) |
|---|---|---|
| Forward Primer | 0.9 | 36 |
| Reverse Primer | 0.9 | 36 |
| Probe | 0.25 | 10 |

**Table SI.3:** Reaction composition for RT-ddPCR assay

| Reagent | Volume (μL) |
|---|---|
| One-step RT-ddPCR supermix | 5.5 |
| Reverse Transcript (10x) | 2.2 |
| 300 mM DTT | 1.1 |
| Primer-Probe mix of each target (40x) | 0.55* |
| RNase/DNase-free water | 0.45 |
| RNA template | 10 |

*Final concentrations in reaction: 0.25μM (probe) and 0.9μM (forward and reverse primers)

**Table SI.4:** Thermal cycling condition for measles, mumps, and rubella virus RT-ddPCR assay

| Cycling Step | Temperature °C | Time | Number of Cycles |
|---|---|---|---|
| Reverse transcription | 50 | 60 min | 1 |
| Enzyme activation | 95 | 10 min | 1 |
| Denaturation | 94 | 30 sec | 40 |
| Annealing/Extension | 55 | 60 sec | |
| Enzyme Deactivation | 98 | 10 min | |
| Hold (optional) | 4 | Infinite | 1 |

**SI 1.4 RT-PCR assays setup and thermal cycling conditions for measles Sanger sequencing**

**Table SI.5:** Primers used for gene amplification of measles with RT-PCR

| Target | Assay Name | Sequence (5'-3') | Accession (coordinates) |
|---|---|---|---|
| L gene (Measles) | Forward Primer | AAAAACGGATTTTCCAACCAAATGA | NC_001498.1 (9393..9417) |
| | Reverse Primer | CTACCAGTGAAGAATACAGGTGTGT | NC_001498.1 (9805..9829) |

| | | |
|---|---|---|
| | Amplicon length | 437 |

**Table SI.6:** Reaction composition for RT-PCR assay

| Reagent | Volume (μL) |
|---|---|
| 2X Platinum™ SuperFi™ RT-PCR Master Mix | 25 |
| Forward primer (10 μM) | 2.5* |
| Reverse primer (10 μM) | 2.5* |
| SuperScript™ IV RT Mix | 0.5 |
| Template RNA | 10 |
| Nuclease-free water | 9.5 |

*Final concentrations in reaction: 0.5μM (forward and reverse primers)

**Table SI.7:** Thermal cycling condition for measles virus RT-PCR assay

| **Cycling Step** | **Temperature** °C | **Time** | **Number of Cycles** |
|---|---|---|---|
| Reverse transcription | 50 | 10 min | 1 |
| RT inactivation/initial denaturation | 98 | 2 min | 1 |
| Amplification | 98 | 10 sec | 40 |
| | 56 | 10 sec | |
| | **72** | 20 sec | |
| Final extension | 72 | 5 min | 1 |
| Hold (optional) | 4 | Infinite | 1 |

**SI 1.5 Calculation of partition coefficients**

**SI. Eq 1:**

$$K_d = \frac{C_S/TSS}{C_w \times (1\,L/1000\,mL)}$$

Where:

$K_d$: partitioning coefficient of target virus in wastewater solid and liquid fraction, mL/g

$C_S$: target viral concentration in wastewater solid fraction, copies/L

$C_W$: target viral concentration in wastewater liquid fraction, copies/L

TSS: wastewater total suspended solids, g/L*

*TSS was measured based on the Standard Method 2540 [7]

### SI 1.6 Calculation of recovery rate and inhibition factor

Recovery rate calculation

**SI. Eq 2:**

$$Recovery\ rate = \frac{C_{WW} \times V_{WW}}{C_{std} \times V_{std}} \times 100\%$$

Where:

$C_{ww}$: concentration of target viral RNA detected in wastewater spiked with virus standards using RT-ddPCR, copies/L

$V_{ww}$: volume of concentrated wastewater sample, L

$C_{std}$: concentration of diluted virus standards spiked into wastewater sample, copies/µL

$V_{std}$: volume of diluted virus standard spiked into each wastewater sample, µL

Inhibition factor calculation

**SI. Eq 3:**

$$Inhibition\ factor = \frac{C_{10-fold}}{C_0}$$

Where:

$C_0$: concentration of target viral RNA detected using original nucleic acid extracts of wastewater spiked with virus standards and RT-ddPCR, copies/L

$C_{10\text{-}fold}$: concentration of target viral RNA detected using 10-fold diluted nucleic acid extracts of wastewater spiked with virus standards and RT-ddPCR, copies/L

**SI 1.7 Measles, mumps, and rubella RNA persistence in wastewater**

**SI. Eq 4:**

$$C_t = C_0 e^{-kt}$$

$$log_{10}\left(\frac{C_t}{C_0}\right) = \frac{-kt}{ln\ (10)}$$

Where:

$C_0$: original viral concentration in the wastewater sample, copies/L

$C_t$: viral concentration in the wastewater sample at day t, copies/L

k: first-order decay constant, 1/days

t: number of days between the measurement of viral concentration in wastewater and the day that the experiment starts, days

**SI. Eq 5:**

$$T_{90} = \frac{ln\ (10)}{k}$$

Where:

$T_{90}$: number of days needed for target viral concentration to decrease by 90% of the original concentration, days

k: first-order decay rate constant, 1/days

**SI 1.8 Quality control measures and limit of detection (LOD) calculation**

The quality control measures and the calculation of LOD are described by Lou et al. (2022)[6] and are summarized briefly as follows. Duplicates of negative control samples were included in the sample treatment, concentration, extraction, and quantification steps to assess potential contamination [8]. Specifically, for wastewater samples spiked with ATCC virus standards, triplicate influent unspiked wastewater samples were stored and processed in the same way and used as negative controls for the treatment. Two 50 mL aliquots of deionized (DI) water were processed in the same way as wastewater samples and used as negative controls for concentration. Two bead tubes containing glass beads and lysis buffer were included as extraction negative controls. The negative controls for concentration and extraction were included in all ddPCR quantification plates containing the wastewater samples that were processed together with the controls. In addition, each ddPCR quantification plate included at least two no-template controls (NTCs) with RNAse-free water and two positive controls using gBlock Gene Fragments (IDT, USA; sequence provided in Tables SI.1).

An acceptable total droplet count of at least 10,000 was established for all sample wells as recommended by the manufacturer. The method limit of detection (LOD) for ddPCR was determined as three positive droplets per well plus the maximum number of positive droplets among the negative controls. The LOD was converted to copies per µL of DNA template and copies per liter of wastewater based on the estimated droplet volume (0.86 nL), the number of total droplets, the volume fraction of DNA template within a droplet (10/22), and the concentration factor during sample processing. The equations used for the LOD calculation are presented in Eq SI. 7-9.

Limit of Detection (LOD) Calculation Equations

**SI. Eq 6:**

$$LOD_{droplet} = 3 + maximum\ number\ of\ positive\ droplets\ across\ all\ process\ blanks^{*}$$

[*] Process blanks include: two concentration blanks, two extraction blanks, and no less than two no template controls per plate included in ddPCR quantification.

**SI. Eq 7:**

$$LOD_{\mu L-DNA\ template}$$

$$= \frac{LOD_{droplet}}{0.86\frac{nl}{droplet} \times n\ total\ droplets\ in\ each\ well^{*} \times \frac{10}{22} fraction\ of\ template\ within\ droplet}$$

$$= \frac{LOD_{droplet} \times 2558.14}{n\ total\ droplets}\ copies\ per\ \mu L\ of\ DNA\ template$$

* The average number of total droplets in each well for measles, mumps, and rubella RT-ddPCR assay is 19884.

**SI. Eq 8:**

$$LOD_{L-wastewater}$$

$$= LOD_{\mu L-DNA\ template} \times 100\ \mu L\ total\ extraction$$

$$\times \frac{1000\ \mu L\ of\ lysis\ buffer\ with\ wastewater\ fraction\ resuspended}{600\ \mu L\ of\ lysis\ buffer\ used\ for\ extraction}$$

$$\times \frac{1}{50\ mL\ wastewater\ concentrated} \times \frac{1000\ mL}{1\ L}$$

$$= LOD_{\mu L-DNA\ template} \times \frac{1000000}{300} copies\ per\ L\ of\ wastewater$$

### SI 2. Results

#### SI 2.1 Measles, mumps and rubella assay validation

**Table SI.8:** Number of mismatches between measles genome sequences and measles assays by genotype

| **Assay** | **Target gene** | **Number of mismatches by genotype** | | | **Source** |
|---|---|---|---|---|---|
| | | WT - D8 | WT - B3 | Edmonston Vaccine | |
| WT1 assay | M gene | 0 | 0 | 1 (probe) | This study |
| WT2 assay | M gene | 0 | 1 (probe) | 2-3 (probe) | This study |
| VA assay | M gene | 2 (probe) | 1 (probe) | 0-1 (probe) | This study |
| CDC WT assay | N gene | 0 | 1 (probe) | 0 | Hummel et al., 2006 [9] |

| | | | | | |
|---|---|---|---|---|---|
| CDC VAC assay | N gene | 1 (probe) | 3 (primers)<br>1 (probe) | 0-1 (primer) | Roy et al., 2017 [10] |

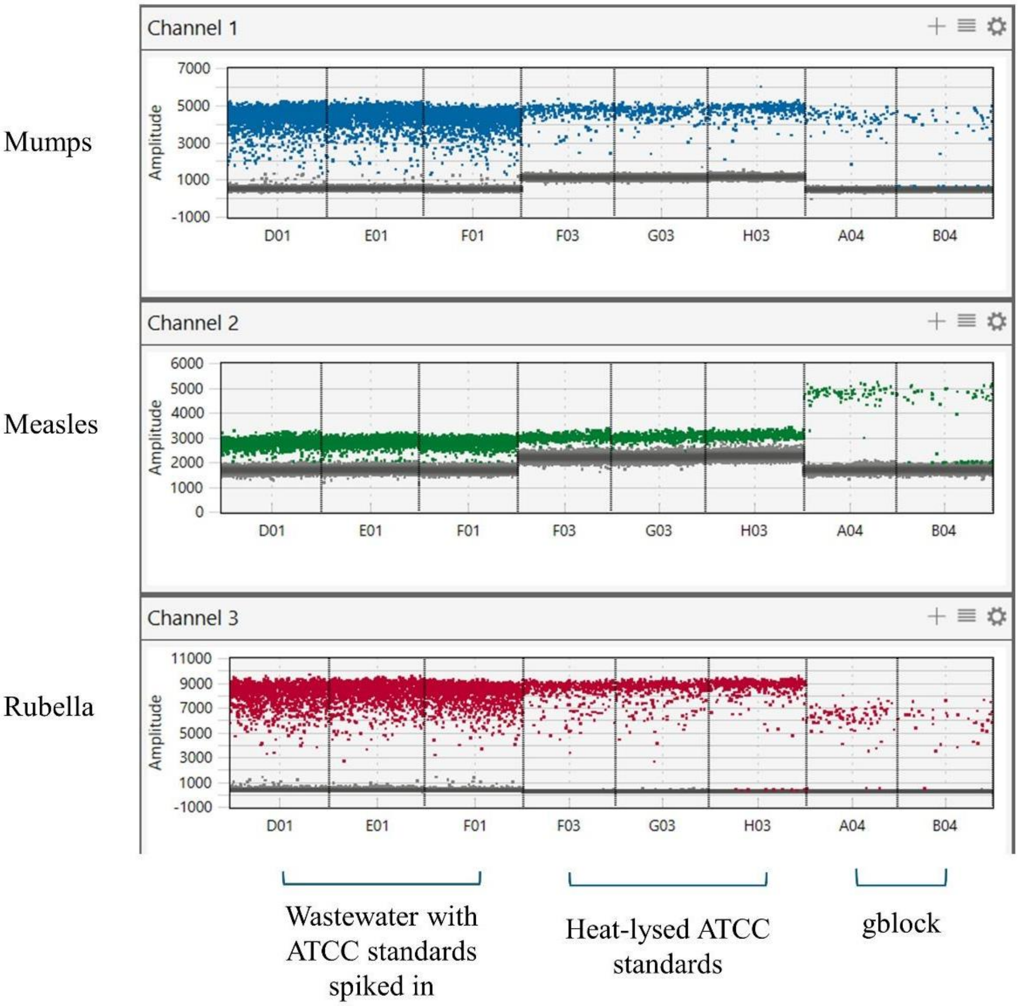

**Figure SI.1.** Multiplexed ddPCR results of measles, mumps, and rubella viral RNA assays using influent wastewater samples spiked with virus standards (columns D01, E01, and F01), heat-lysed virus standards (F03, G03, and H03), and gblock standards (A04, B04).

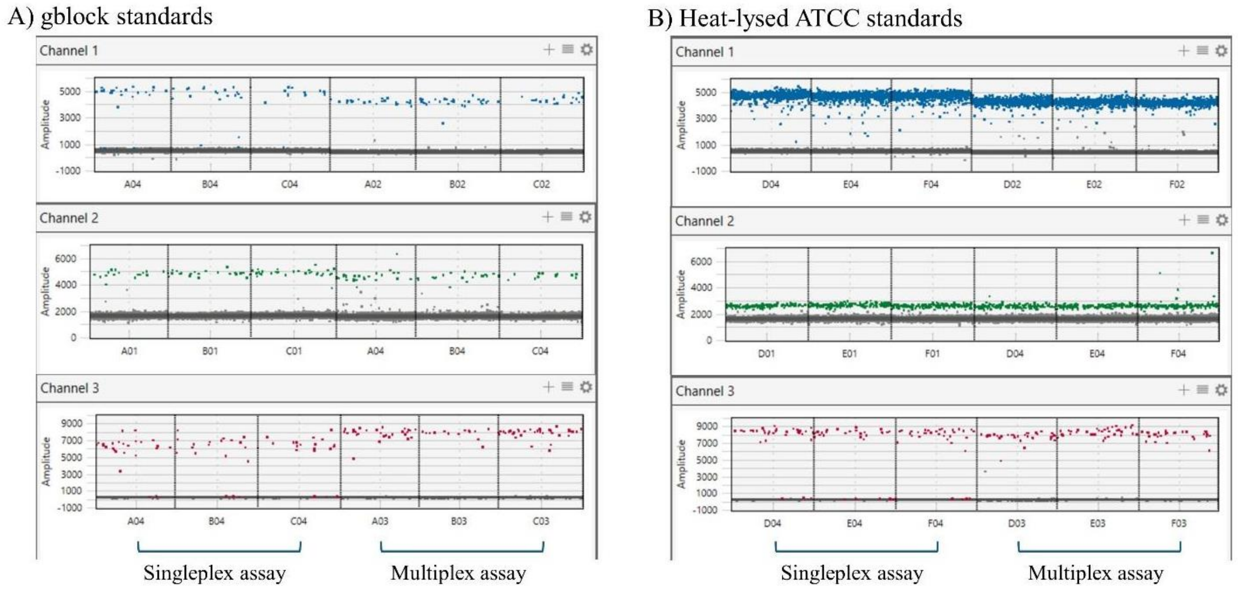

**Figure SI.2.** Comparison of singleplex and multiplex RT-ddPCR quantification of measles, mumps, and rubella viral RNA using gblock standards (A) and heat-lysed virus standards (B).

**Table SI.9:** Concentration of measles, mumps, and rubella viral RNA in gblock standards and heat-lysed virus standards measured with singleplex and multiplex RT-ddPCR assays.

| Target | Type of standards | Singleplex concentration (± standard error, copies/µL) | Multiplex concentration (± standard error, copies/µL) | p-value |
|---|---|---|---|---|
| Measles | gblock | 3.37 ± 0.37 | 2.70 ± 0.47 | 0.33 |
| Mumps | gblock | 3.19 ± 0.37 | 3.03 ± 0.68 | 0.85 |
| Rubella | gblock | 3.67 ± 0.78 | 3.16 ± 0.35 | 0.60 |
| Measles | Heat-lysed virus | 19.19 ± 0.29 | 20.52 ± 0.92 | 0.29 |
| Mumps | Heat-lysed virus | 105.96 ± 3.92 | 98.81 ± 0.27 | 0.21 |
| Rubella | Heat-lysed virus | 4.63 ± 0.11 | 4.63 ± 0.20 | 0.99 |

**Table SI.10:** Concentrations of wild type B3 and vaccine measles strains measured with measles WT2 and VA singleplex and multiplex RT-ddPCR assays.

| Strain of standards | Concentration measured with WT2 assay (± standard error, log 10 (copies/L-wastewater) | Concentration measured with VA assay (± standard error, log 10 (copies/L-wastewater) |
|---|---|---|

| | | |
|---|---|---|
| B3 and vaccine[1] | 5.41 ± 0.06 | 5.43 ± 0.06 |
| B3 and vaccine[2] | 4.22 ± 0.09 | 5.39 ± 0.06 |
| B3 | 4.28 ± 0.03 | 4.27 ± 0.03 |
| Vaccine | 5.36 ± 0.02 | 5.36 ± 0.02 |

[1] Both assays measured the concentration of both measles strains (wild type B3 and vaccine strain)

[2] The WT2 assay only measured the concentration of wild type B3 measles strain, and the VA assay only measured the concentration of vaccine measles strain. The droplets from ddPCR were distinguished using different amplitudes.

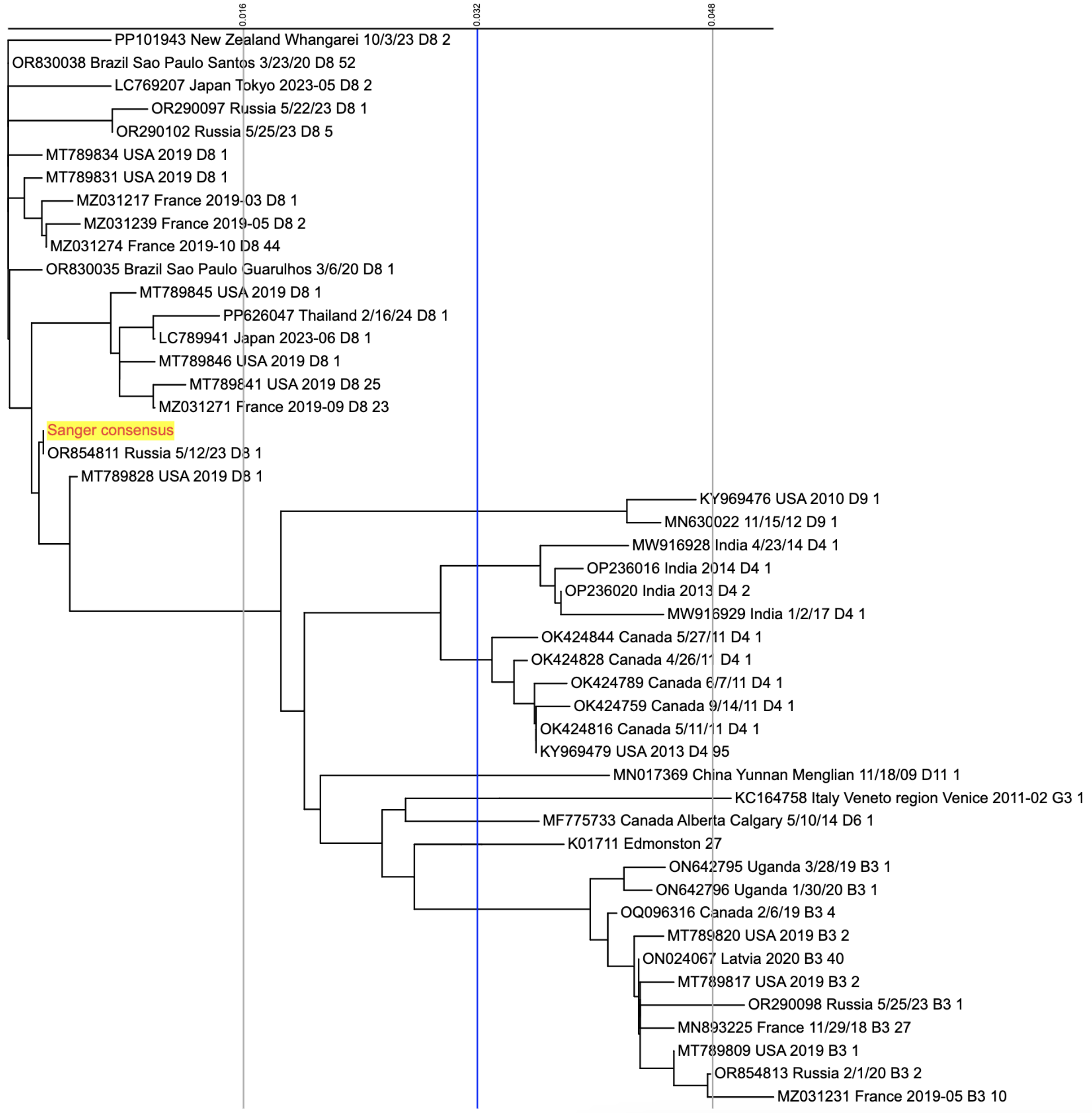

**Figure SI.3.** A phylogenetic tree based on the consensus sequence of the Sanger sequencing replicates of selected measles-positive wastewater samples. The phylogenetic tree was generated using iTOL. The positive wastewater sample contained D8 genotype measles virus.

**SI 2.2 Evaluating recovery of measles, mumps, and rubella RNA from liquid and solid fractions of influent wastewater samples**

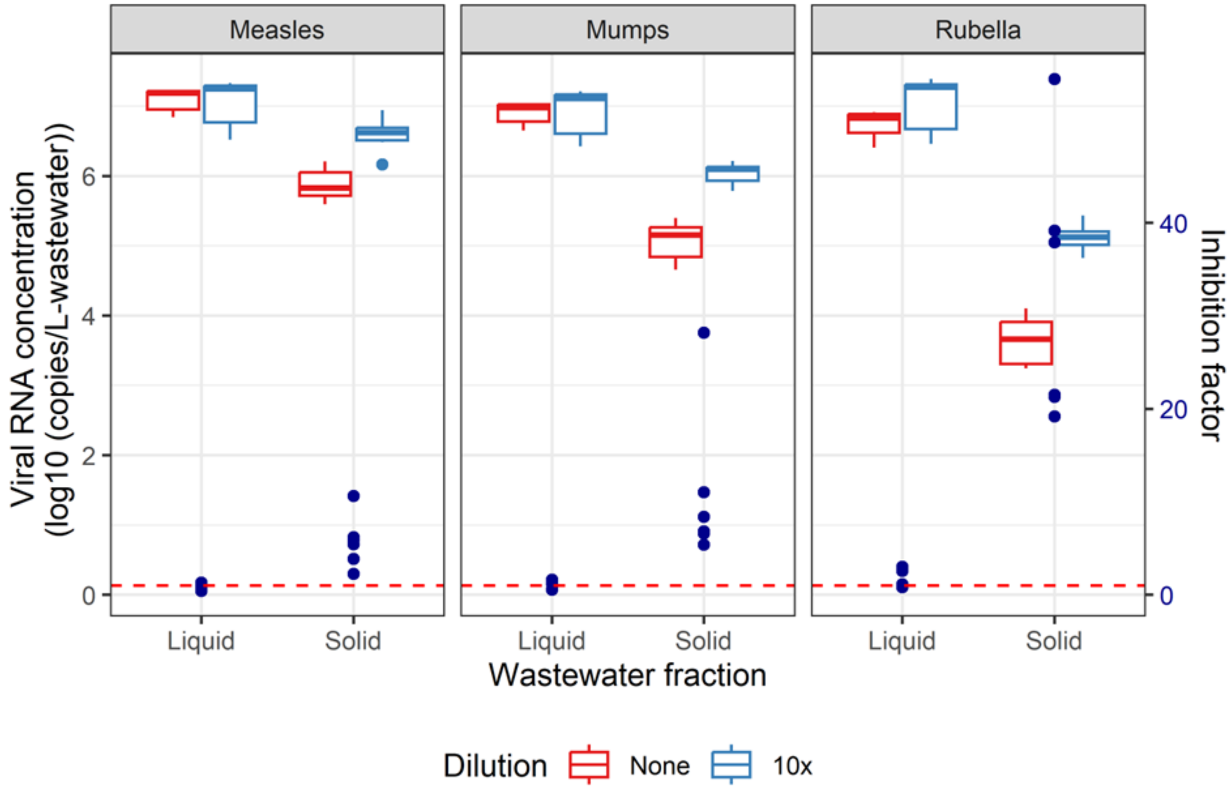

**Figure SI.4.** Boxplots of measles, mumps, and rubella viral RNA concentrations in influent wastewater liquid and solid fractions. Concentrations were determined using undiluted (red) and ten-fold diluted (blue) nucleic acid extracts from wastewater samples spiked with virus standards. Dark blue dots indicated the inhibition factor (calculations as described in section SI

1.3). Higher inhibition factors indicate greater inhibition of RT-ddPCR. The red dashed line indicates no inhibition (inhibition factor = 1).

**SI 2.3 Measles, mumps, and rubella RNA persistence in wastewater**

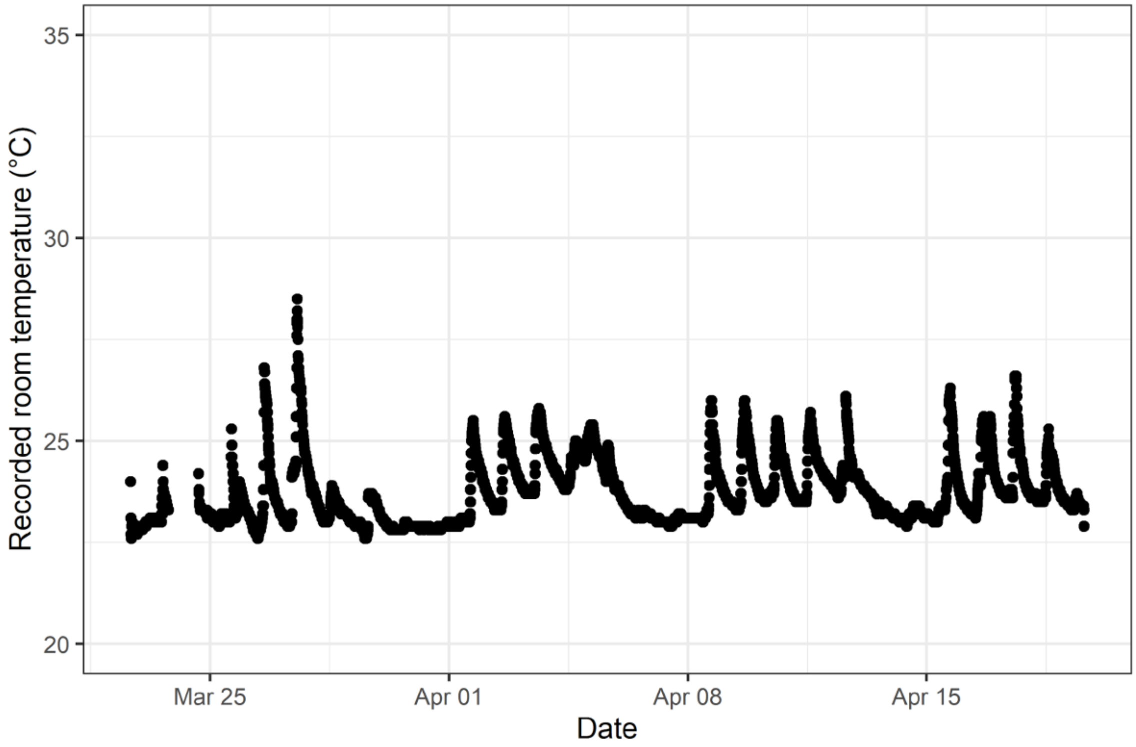

**Figure SI.5.** Logged room temperature measurements during the persistence experiment. Temperature data on day 2 is missing due to an issue with the temperature measurement instrument.

**Table SI.11:** Results from measles, mumps, and rubella RNA persistence tests in wastewater. Initial RNA concentrations (day = 0) of each target in each condition from triplicates are shown. The results include the first-order decay constant (k) with standard errors, the significance level of the first-order decay constant (p-value), the number of days required for 90% reduction ($T_{90}$), and the $R^2$ of the linear regression.

| Target | Log-transformed concentration at day 0 (copies/L-wastewater) | Temperature | k (± standard error) | p-value | $T_{90}$ | $R^2$ |
|---|---|---|---|---|---|---|
| Measles | 7.09 ± 0.18 | 4°C | 0.057 ± 0.008 | <0.001 | 40.5 | 0.74 |
| | | Room | 0.282 ± 0.017 | <0.001 | 8.2 | 0.94 |
| | 6.17 ± 0.03 | 4°C | 0.079 ± 0.004 | <0.001 | 29.3 | 0.96 |
| | | Room | 0.369 ± 0.014 | <0.001 | 6.2 | 0.98 |
| Mumps | 6.90 ± 0.17 | 4°C | 0.050 ± 0.007 | <0.001 | 45.7 | 0.72 |
| | | Room | 0.205 ± 0.011 | <0.001 | 11.2 | 0.95 |
| | 6.00 ± 0.06 | 4°C | 0.060 ± 0.005 | <0.001 | 38.4 | 0.88 |
| | | Room | 0.253 ± 0.010 | <0.001 | 9.1 | 0.97 |
| Rubella | 6.74 ± 0.21 | 4°C | 0.075 ± 0.007 | <0.001 | 30.8 | 0.84 |
| | | Room | 0.441 ± 0.024 | <0.001 | 5.2 | 0.95 |
| | 5.86 ± 0.06 | 4°C | 0.073 ± 0.004 | <0.001 | 31.6 | 0.95 |
| | | Room | 0.452 ± 0.024 | <0.001 | 5.1 | 0.96 |

### References

1 E. W. Sayers, E. E. Bolton, J. R. Brister, K. Canese, J. Chan, D. C. Comeau, R. Connor, K. Funk, C. Kelly, S. Kim, T. Madej, A. Marchler-Bauer, C. Lanczycki, S. Lathrop, Z. Lu, F. Thibaud-Nissen, T. Murphy, L. Phan, Y. Skripchenko, T. Tse, J. Wang, R. Williams, B. W. Trawick, K. D. Pruitt and S. T. Sherry, Database resources of the national center for biotechnology information, *Nucleic Acids Res.*, 2022, **50**, D20–D26.

2 K. Katoh, J. Rozewicki and K. D. Yamada, MAFFT online service: multiple sequence alignment, interactive sequence choice and visualization, *Brief. Bioinform.*, 2019, **20**, 1160–1166.

3 A. Untergasser, I. Cutcutache, T. Koressaar, J. Ye, B. C. Faircloth, M. Remm and S. G. Rozen, Primer3—new capabilities and interfaces, *Nucleic Acids Res.*, 2012, **40**, e115.

4 C. Camacho, G. Coulouris, V. Avagyan, N. Ma, J. Papadopoulos, K. Bealer and T. L. Madden, BLAST+: architecture and applications, *BMC Bioinformatics*, 2009, **10**, 421.

5 Z. W. Laturner, D. M. Zong, P. Kalvapalle, K. R. Gamas, A. Terwilliger, T. Crosby, P. Ali, V. Avadhanula, H. H. Santos, K. Weesner, L. Hopkins, P. A. Piedra, A. W. Maresso and L. B. Stadler, Evaluating recovery, cost, and throughput of different concentration methods for SARS-CoV-2 wastewater-based epidemiology, *Water Res.*, 2021, **197**, 117043.

6 E. G. Lou, N. Sapoval, C. McCall, L. Bauhs, R. Carlson-Stadler, P. Kalvapalle, Y. Lai, K. Palmer, R. Penn, W. Rich, M. Wolken, P. Brown, K. B. Ensor, L. Hopkins, T. J. Treangen and L. B. Stadler, Direct comparison of RT-ddPCR and targeted amplicon

sequencing for SARS-CoV-2 mutation monitoring in wastewater, *Sci. Total Environ.*, 2022, **833**, 155059.

7 In *Standard Methods For the Examination of Water and Wastewater*, American Public Health Association, 2017.

8 M. A. Borchardt, A. B. Boehm, M. Salit, S. K. Spencer, K. R. Wigginton and R. T. Noble, The Environmental Microbiology Minimum Information (EMMI) Guidelines: qPCR and dPCR Quality and Reporting for Environmental Microbiology, *Environ. Sci. Technol.*, 2021, **55**, 10210–10223.

9 K. B. Hummel, L. Lowe, W. J. Bellini and P. A. Rota, Development of quantitative gene-specific real-time RT-PCR assays for the detection of measles virus in clinical specimens, *J. Virol. Methods*, 2006, **132**, 166–173.

10 F. Roy, L. Mendoza, J. Hiebert, R. J. McNall, B. Bankamp, S. Connolly, A. Lüdde, N. Friedrich, A. Mankertz, P. A. Rota and A. Severini, Rapid Identification of Measles Virus Vaccine Genotype by Real-Time PCR, *J. Clin. Microbiol.*, 2017, **55**, 735–743.
